# Private Equity Practice Connections and Medicare Professional Payments Among US Neurological Surgeons

**DOI:** 10.64898/2026.11.13.26368057

**Authors:** Abhinav Bachu, Raghav Rajesh, Zachary Uram, Anoop Chinthala, Anand V. Germanwala

**Affiliations:** Department of Neurological Surgery, Loyola University Chicago Stritch School of Medicine, 2160 S. First Avenue, Maywood, IL 60153, United States

**Keywords:** neurological surgery, private equity, Medicare, physician practice, management services organization

## Abstract

**Background:** Medicare payment patterns among neurosurgeons with private equity connections remain poorly characterized. We compared annual professional payments by connection type.

**Methods:** In this retrospective cross-sectional study, public Medicare provider summaries for 2013–2024 were linked to dated affiliation and investment records. The primary comparison included 69 practice or management-services organization–connected surgeons and 4,016 comparison surgeons. Connected surgeons contributed index-year payments; comparison surgeons contributed averages over eligible years. Welch comparisons of log payments estimated geometric mean ratios. Supporting analyses examined dollar differences, state-year standardization, beneficiary age/risk balance, 35 hospital-operator surgeons, and longitudinal trajectories. Payments were expressed in 2024 US dollars.

**Results:** The primary geometric mean ratio was 1.38 (95% confidence interval 1.11–1.72; P = .004). The arithmetic mean difference was +$31,250 (−$3,006 to +$65,506); the state-year standardized difference was +$32,204 (+$2,886 to +$65,372). Age/risk balancing attenuated the ratio to 1.25 (1.01–1.59). The hospital-operator ratio was 0.53 (0.35–0.81); the pooled ratio was 1.00 (0.81–1.25). Among 50 longitudinally eligible practice or management-services organization–connected surgeons, the differential annual change was +16.9% (−0.3% to +37.2%) on the log scale and −$15,727 (wild cluster bootstrap interval −$95,893 to +$35,191) in dollars. Excluding one high-billing surgeon reversed the dollar estimate. All intervals are 95% confidence intervals.

**Conclusion:** Practice and management-services organization connections were associated with higher unadjusted payments; balancing attenuated the ratio to 1.25. Hospital-operator payments were lower. Longitudinal estimates were imprecise and do not establish acquisition effects.

## Introduction

Private equity (PE) investment has become an increasingly visible feature of physician practice organization, including surgical specialties.[10] Practices may join investor-backed platforms through direct acquisition, management-services arrangements, or other economic relationships. These arrangements can affect referral patterns and how clinical services are scheduled and delivered. Studies in other procedural specialties have linked PE involvement to differences in spending and utilization, although their findings vary by specialty and outcome.[2,5,8,11] For neurological surgery, comparing the professional activity of surgeons in investor-linked practices with that of other surgeons can inform discussions of practice organization and consolidation.

The neurosurgical literature has examined the future of private practice and the implications of PE investment.[1] Recent conference abstracts have also described PE-backed neurosurgical care and consolidation in spine ambulatory surgery centers.[6,7] Physician-level payment comparisons extend this transaction-focused literature by examining the professional activity of neurosurgeons with documented investment connections. A practice or management-services organization (MSO) relationship is particularly relevant to this question because it links investment to the organization of physician services. Surgeons employed by investor-owned hospital systems may have different payment patterns from those in PE-backed physician practices.

Medicare provider summaries offer a consistent national source of annual professional payments and service measures.[3] These payments reflect the volume and composition of reimbursed care, rather than physician compensation alone. Traditional Medicare rates generally follow an administered fee schedule, while annual provider payments can vary with beneficiary volume, service mix, geography, and billing. We compared annual Medicare professional payments among neurosurgeons with documented PE-backed practice or practice-linked MSO connections and comparison surgeons without an identified connection. The main comparison examined proportional payment differences using geometric means. Supporting analyses examined absolute dollar differences, geographical and temporal composition, and payment trajectories around organizational investment. Hospital-operator relationships and the pooled investment-connected cohort were evaluated separately to place the practice/MSO findings in context.

## Materials and Methods

### Study design and data sources

We conducted a retrospective observational study using Centers for Medicare & Medicaid Services (CMS) Medicare Physician & Other Practitioners provider summaries for 2013–2024. [3] The study was a descriptive physician-level cross-sectional comparison, centered on practice/MSO connections. Connected surgeons contributed one index-year observation, and comparison surgeons contributed their mean payment over eligible years. A supporting longitudinal analysis used annual records around the start of the organization’s recorded investment window. The connected index observations occurred in 2015 and 2018–2024 across 30 states and 52 state-year strata. This study is reported in accordance with the Strengthening the Reporting of Observational Studies in Epidemiology (STROBE) guideline.

Potential PE-backed organizations were identified using PitchBook and targeted searches of public company announcements and investment-firm websites through October 2026. Physician affiliations were established from dated practice rosters and archived records and linked to CMS data by National Provider Identifier (NPI). One investigator reviewed the supporting evidence to confirm professional relationships and investment timing. Dated CMS group-membership records and other professional-affiliation documents supplemented the roster review. Supplementary Methods and Table S1 describe the classification and organization-specific analytical windows.

### Connected and comparison cohorts

Inclusion required an individual CMS provider record classified as neurosurgery, a dated qualifying professional relationship, and an organizational investment link covering the dated observation. Qualifying relationships included membership in a PE-backed practice, a practice-linked MSO arrangement, or employment in a hospital-operator professional group. Documented minority equity investment qualified; hospital privileges alone and debt-only financing did not. Unresolved relationships and observations outside the supported window were not included. After duplicate removal, each surgeon’s earliest eligible dated observation with same-year CMS presence defined the index year.

The main exposure group comprised 69 practice/MSO-connected surgeons. A separate group of 35 hospital-operator professional-group surgeons provided a secondary comparison. The latter category included LifePoint, Steward, and Quorum, encompassing both conventional PE sponsorship and investment-fund ownership following restructuring. This broader employment category was analyzed separately from practice/MSO relationships. BASS/Privia was classified through its practice-linked MSO and retained sponsor equity during the 2022 index observations; public listing alone was not an exclusion. These definitions identify professional economic connections and do not imply that every practice was directly acquired.

Comparison surgeons were individual neurosurgeons with provider summaries in the connected cohort’s state-year strata. Physicians with known or unresolved professional PE connections in the screening inventory, and specified exact organizational matches, were excluded from the comparison pool across years. The resulting pool contained 4,016 distinct physicians and 8,118 physician-years. Connected and comparison NPIs did not overlap. Comparisons were geographically and temporally restricted, rather than individually matched; the term comparison denotes no identified connection within the reviewed inventory.

### Outcomes

The primary outcome was annual provider-wide Medicare professional payment, CMS Tot_Mdcr_Pymt_Amt, converted to 2024 US dollars using the annual all-items Consumer Price Index for All Urban Consumers.[13] Payments represent traditional Medicare fee-for-service activity attributed to the provider and exclude facility revenue, Medicare Advantage payments, and commercial insurance activity. Annual beneficiary counts and service units provided practice context. Beneficiaries were distinct within physician-year, and service units were billed units rather than surgical cases. Provider-level mean beneficiary age and risk score were used in a supporting balancing analysis. Missing or suppressed values were not assigned zero; payment, age, and risk variables were complete in the cross-sectional analytical records.

### Statistical analysis

The main analysis compared natural-log annual payments between practice/MSO-connected surgeons and comparison surgeons using a two-sided Welch t-test. Practice/MSO connections identify the physician-practice investment exposure of interest, and the log scale summarizes proportional payment differences while reducing the influence of extreme dollar values in the right-skewed distribution. Exponentiating the difference in mean log payments and its 95% confidence interval (CI) yielded the geometric mean ratio. Each practice/MSO surgeon contributed an index-year payment; each comparison surgeon contributed the arithmetic mean of eligible annual payments before log transformation. Physicians were weighted equally within the group.

Supporting analyses compared arithmetic mean payments with Welch CIs, allowing unequal group variances, and median payments with physician-level bootstrap CIs. Mann–Whitney tests assessed distributional differences, not equality of medians specifically. The same dollar and log comparisons were reported separately for hospital-operator connections and for the pooled 104-surgeon connected cohort. All estimates and P values were nominal, without multiplicity adjustment, and were interpreted together as descriptive observational comparisons.

State-year standardization weighted each within-stratum arithmetic mean difference by that stratum’s share of connected surgeons, using the practice/MSO distribution for the practice/MSO estimate. Relative-entropy weighting was applied separately to the practice/MSO and pooled cohorts, preserving each cohort’s state-year totals while balancing mean provider-level beneficiary age and risk score. Practice/MSO weighting estimated dollar differences and geometric mean ratios from annual payment records; the state-year-only log contrast used the same annual-record aggregation. These weighted contrasts target a different composition from the main comparison of physician-level payment summaries. Percentile CIs used 19,999 bootstrap draws for practice/MSO-balanced estimates and the state-year-only log ratio, and 4,999 for the other standardized, pooled-balanced, and median estimates. Draws resampled physicians within each group, retained all eligible annual records and multiplicities, and refitted weights. The practice/MSO annual-record bootstrap sampled 69 connected surgeons and the 3,294 comparison physicians contributing records in its 36 eligible state-year strata. These cross-sectional procedures assume independence between physicians and do not adjust for shared organizations.

The supporting longitudinal analysis included 50 practice/MSO surgeons and 20 hospital-operator surgeons with at least one CMS record in the index state before organizational investment and at least one in the following two years. A stacked difference-in-differences model used state–investment-year subexperiments, physician-by-subexperiment and year-by-subexperiment fixed effects, and comparison sample-share weights.[14] The investment year was excluded. Standard errors were clustered by connected parent organization and by comparison physician, using a t reference with connected-organization clusters minus one degrees of freedom. Dollar, log-payment, and event-study models characterized payment trajectories.

Sensitivity analyses stratified cross-sectional comparisons by provider-address Rural–Urban Commuting Area (RUCA) codes (1–3 metropolitan; 4–10 nonmetropolitan), omitted influential parent organizations, and excluded uncertain affiliation index years. For practice/MSO longitudinal models, we used a restricted wild cluster bootstrap t CI (9,999 Webb six-point draws), omitted each parent in turn, removed surgeons above the 99th and 95th percentiles of preinvestment mean payment, and restricted the cohort to 46 surgeons with a preinvestment record matching their index ZIP code. Hospital-operator DiD results were reported as point estimates because only three connected parent clusters were available. Organizational investment dates, rather than verified individual acquisition dates, anchored the longitudinal models. Detailed methods and results are in the supplement. Analyses used Python 3.12.14, NumPy 2.3.5, and SciPy 1.17.0. Two-sided P < .05 was the nominal significance threshold.

### Ethics

Institutional review board review was not required for this study, which used publicly available provider-level records and contained no individual patient clinical records.

## Results

### Cohort characteristics

The source inventory contained 10,474 candidate records: 3,046 accepted, 2,130 rejected, and 5,298 unresolved. Removing 34 duplicate accepted records yielded 3,012 eligible dated professional observations representing 104 distinct connected surgeons (Figure 1). These source-record counts reflect documentary observations, not unique physicians. The cohort comprised 69 practice/MSO surgeons across 20 parent-organization labels and 35 hospital-operator surgeons across three labels (Table 1). LifePoint accounted for 27 hospital-operator surgeons, Steward for five, and Quorum for three. Practice/MSO and comparison surgeons had mean beneficiary ages of 73.2 and 71.8 years and risk scores of 1.30 and 1.53, respectively.

**Table 1:** Cohort and beneficiary characteristics.

| **Characteristic** | **Practice/MSO** | **Hospital operator** | **Comparison pool** |
|---|---|---|---|
| Distinct physicians | 69 | 35 | 4,016 |
| Contributing physician-years | 69 | 35 | 8,118 |
| Provider states | 22 | 18 | 30 |
| Represented state-year strata | 36 | 23 | 52 |
| Connected parent-organization labels | 20 | 3 | — |
| Index calendar years | 2015 and 2018–2023 | 2019–2024 | Eligible connected state-years |
| Mean beneficiary age in years | 73.2 | 72.0 | 71.8 |
| Mean beneficiary risk score | 1.30 | 1.53 | 1.53 |
| Missing payment, age, or risk records | 0 | 0 | 0 |

The main exposure group comprises practice/MSO surgeons. Hospital-operator surgeons are evaluated separately; their union forms the 104-surgeon pooled cohort. State and state-year counts overlap between connection types. Comparison surgeons are drawn from the 52 state-year strata represented by the full documented-connection inventory.
Age and risk are averages of provider-level beneficiary means, with each physician weighted equally. Comparison physicians first contribute their mean over eligible years. In the pooled 104-surgeon connected cohort, mean beneficiary age is 72.8 years and risk score is 1.38; the corresponding state-year standardized comparison means are 71.8 years and 1.50. These standardized comparison values differ from the equally weighted physician means displayed in this table.

**Figure 1:**
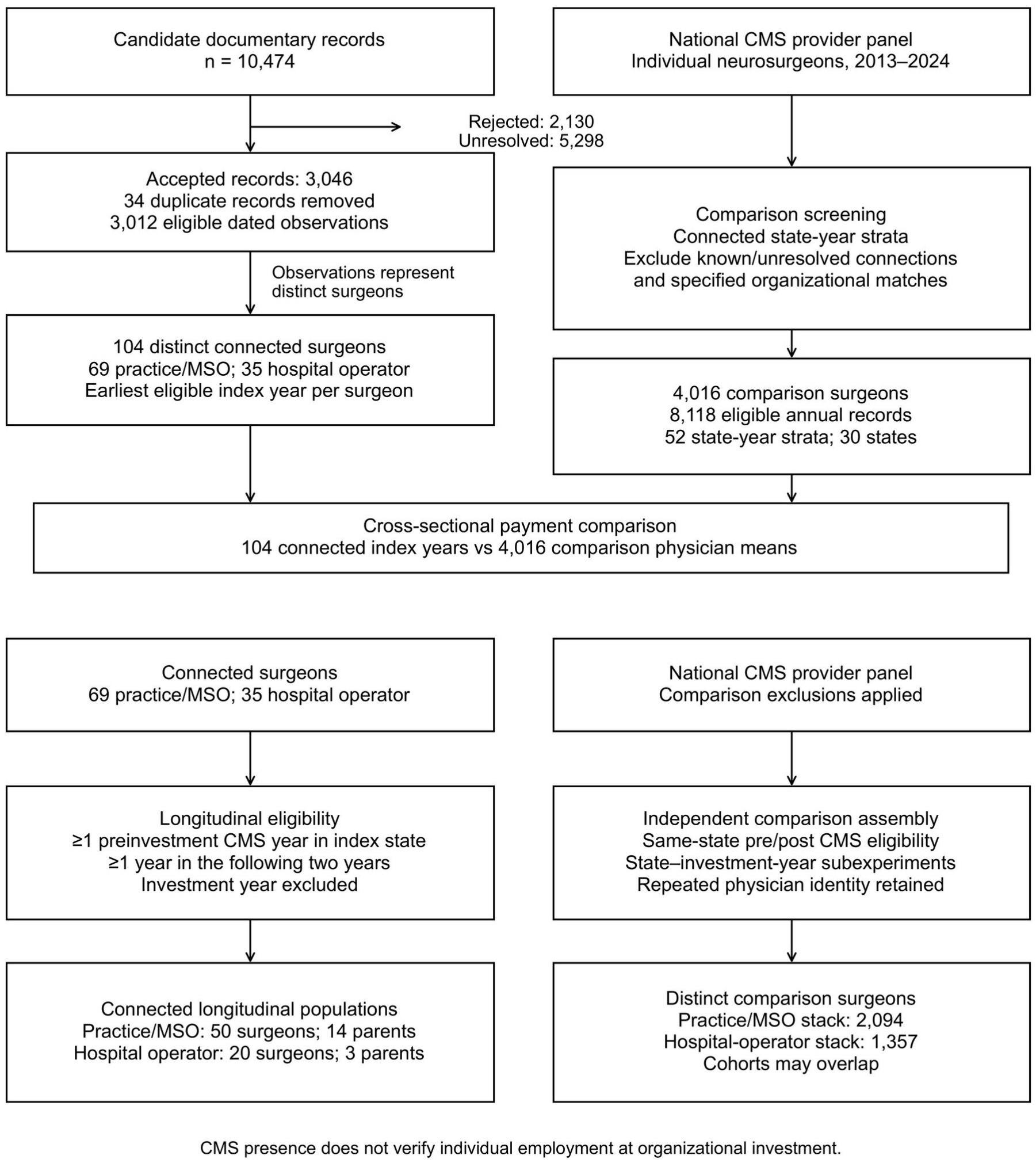
Source flow and analytical populations. A, Candidate documentary records yielded 3,012 eligible dated observations representing 104 connected surgeons after duplicate removal. Connected surgeons contributed their earliest eligible index year; 4,016 comparison surgeons contributed their mean over eligible annual records in the connected cohort’s state-year strata. B, Longitudinal eligibility required a CMS record in the index state before organizational investment and in at least one of the following two years, excluding the investment year. Practice/MSO and hospital-operator populations included 50 and 20 connected surgeons, respectively. Comparison cohorts were assembled independently and may overlap. Documentary records, distinct surgeons, and repeated subexperiment memberships are different denominators. CMS presence does not verify employment at investment start. CMS, Centers for Medicare & Medicaid Services; MSO, management-services organization. Alt text: Two-panel flow distinguishes 10,474 documentary records from 104 connected surgeons and 4,016 cross-sectional comparison surgeons. Both branches feed the payment comparison. Independently assembled longitudinal analyses include 50 practice/MSO and 20 hospital-operator surgeons, with 2,094 and 1,357 distinct comparison surgeons.

#### Practice and MSO payment comparisons

Practice/MSO-connected surgeons had geometric mean annual Medicare professional payments of $114,555, compared with $82,829 among comparison surgeons. The geometric mean ratio was 1.38 (95% CI 1.11–1.72; P = .004), corresponding to 38% higher geometric mean payments (Table 2; Figure 2). Arithmetic mean payments were $163,087 and $131,837, respectively, with a difference of +$31,250 (95% CI −$3,006 to +$65,506; P = .073). Median payments were $117,101 and $98,094; the median difference was +$19,007 (bootstrap 95% CI −$5,406 to +$70,125; Table S2).

**Table 2:** Physician-level annual Medicare professional payment comparisons.

| **Connected group and scale** | **n** | **Contrast** | **95% CI** | **P** |
|---|---|---|---|---|
| Practice/MSO<br>Log payment | 69 | Ratio 1.38 | 1.11 to 1.72 | .004 |
| Practice/MSO<br>Dollar payment | 69 | +\$31,250 | −\$3,006 to +\$65,506 | .073 |
| Hospital operator<br>Log payment | 35 | Ratio 0.53 | 0.35 to 0.81 | .004 |
| Hospital operator<br>Dollar payment | 35 | −\$46,328 | −\$88,708 to −\$3,949 | .033 |
| All connected<br>Log payment | 104 | Ratio 1.00 | 0.81 to 1.25 | .987 |
| All connected<br>Dollar payment | 104 | +\$5,142 | −\$22,362 to +\$32,646 | .712 |

Dollar contrasts are connected-minus-comparison arithmetic mean differences in 2024 US dollars. Log contrasts are geometric mean ratios. Comparison arithmetic and geometric means are \$131,837 and \$82,829 among 4,016 physicians. Welch intervals allow unequal variances and assume independent physicians. Each connected surgeon contributes an index year; each comparison surgeon contributes the arithmetic mean of eligible years before log transformation. P values are nominal and unadjusted for multiple comparisons.

**Figure 2:**
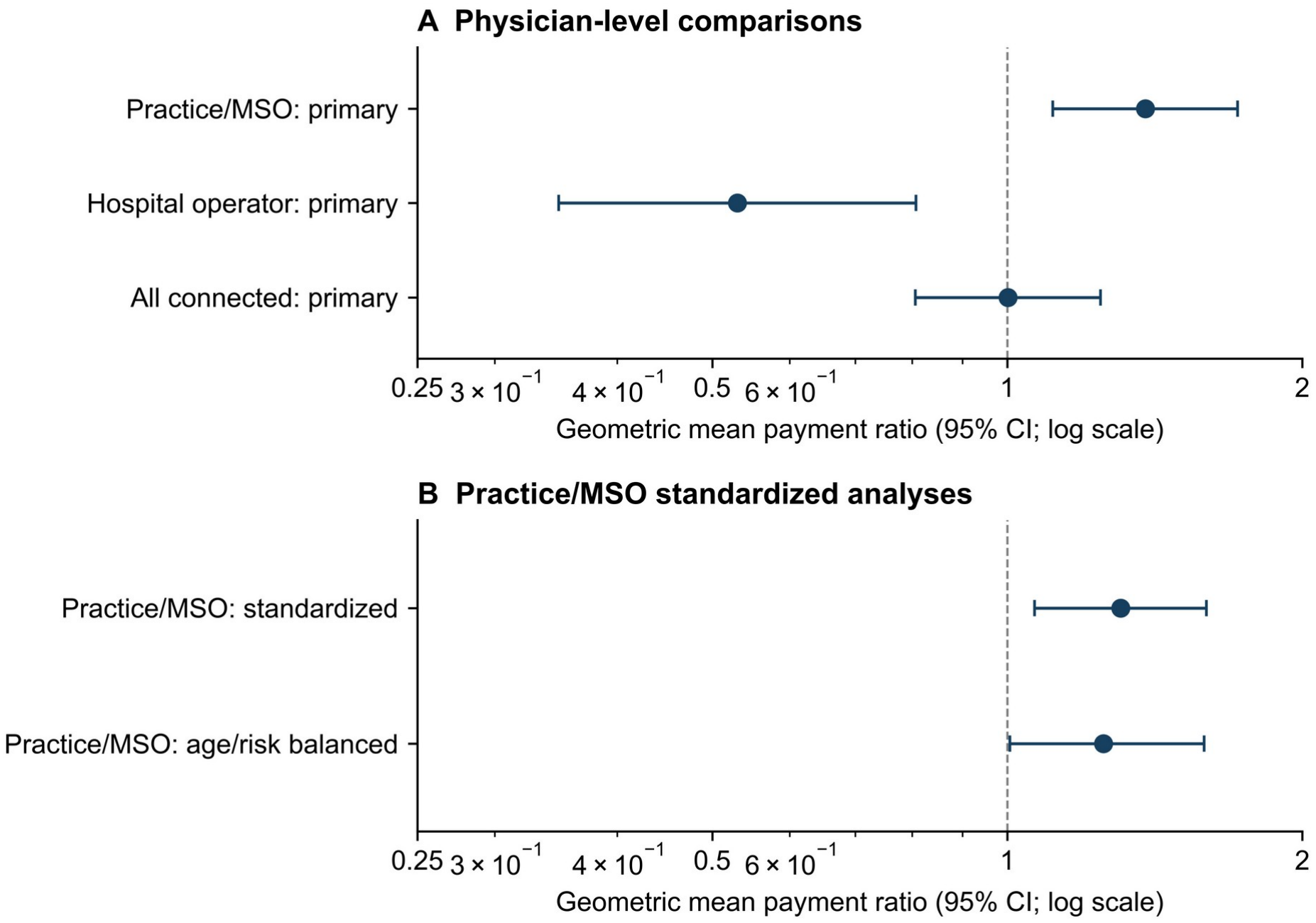
Geometric mean Medicare professional payment ratios. A, Geometric mean payment ratios and Welch 95% confidence intervals (CIs) compare practice/MSO surgeons (n = 69), hospital-operator surgeons (n = 35), and their pooled cohort (n = 104) with the same 4,016 comparison physicians. Connected surgeons contribute index-year payments; comparison physicians contribute the arithmetic mean of eligible annual payments before log transformation. B, Practice/MSO contrasts use annual log-payment records and the connected state-year distribution. Relative-entropy weighting additionally balances mean provider-level beneficiary age and risk score. The comparison population contains 3,294 physicians contributing 5,943 eligible annual records in 36 strata. Percentile CIs use 19,999 physician-level bootstrap draws from these eligible comparison physicians and 69 connected surgeons, retaining annual-record multiplicities and refitting weights. The panels represent distinct estimands, not successive adjustments to the same physician-summary outcome. Points show ratios; bars show 95% CIs; the logarithmic axis has a null value of 1. Balancing attenuates the practice/MSO point estimate to approximately 25% higher payments. Intervals are nominal and do not account for shared organizations. MSO, management-services organization. Alt text: Two forest panels distinguish connection-type comparisons from weighted annual-record practice/MSO analyses. Practice/MSO payments are higher and hospital-operator payments lower than comparison payments; the pooled ratio is near one. Beneficiary age/risk balancing attenuates the practice/MSO point estimate to 1.25, with a nominal 95% CI of 1.01– 1.59.

The practice/MSO state-year standardized arithmetic mean difference was +$32,204 (95% CI +$2,886 to +$65,372; Table 3). After age/risk balancing, the dollar difference was +$19,708 (95% CI −$10,419 to +$52,919) and the geometric mean ratio was 1.25 (95% CI 1.01–1.59). The corresponding state-year-only log ratio was 1.31 (95% CI 1.07–1.60; Figure 2B). Findings persisted among metropolitan providers and after omitting influential parent organizations or uncertain index-year assignments (Tables S5–S9).

**Table 3:** Standardized and beneficiary-balanced payment comparisons.

| **Analysis and scale** | **Contrast** | **Bootstrap 95% CI** |
|---|---|---|
| Practice/MSO state-year standardized<br>Dollar difference | +\$32,204 | +\$2,886 to +\$65,372 |
| Practice/MSO state-year standardized<br>Geometric mean ratio | Ratio 1.31 | 1.07 to 1.60 |
| Practice/MSO with age/risk balance<br>Dollar difference | +\$19,708 | −\$10,419 to +\$52,919 |
| Practice/MSO with age/risk balance<br>Geometric mean ratio | Ratio 1.25 | 1.01 to 1.59 |
| Hospital operator state-year standardized<br>Dollar difference | −\$50,464 | −\$83,099 to −\$8,807 |
| All connected state-year standardized<br>Dollar difference | +\$4,383 | −\$20,531 to +\$32,291 |
| All connected with age/risk balance<br>Dollar difference | −\$4,900 | −\$29,306 to +\$22,584 |

Dollar contrasts are differences in arithmetic mean annual payments in 2024 US dollars. Ratios compare geometric means. Standardization uses each connected group’s state-year distribution; age/risk balance additionally matches that group’s mean provider-level beneficiary age and risk score. Weighted comparisons use annual comparison records. Their log ratios therefore differ in aggregation and weighting from the main physician-summary ratio in Table 2.
Practice/MSO weights cover 5,943 annual comparison records from 3,294 physicians in 36 state-year strata, with target age 73.2 years and risk score 1.30. Intervals use 19,999 physician-level bootstrap draws for practice/MSO-balanced contrasts and the state-year-only log ratio; other rows use 4,999 draws. Weights are refitted in every draw. Intervals do not account for shared organizations. The balanced ratio CI is 1.01–1.59, with a lower endpoint close to unity. Bootstrap intervals have finite Monte Carlo precision.

#### Hospital-operator and pooled comparisons

Hospital-operator surgeons had lower payment levels, with a geometric mean ratio of 0.53 (95% CI 0.35–0.81) relative to comparison surgeons. Their arithmetic mean payment was $85,508, with a difference of −$46,328 (95% CI −$88,708 to −$3,949; P = .033), and their median payment was $58,678. The state-year standardized mean difference was −$50,464 (95% CI −$83,099 to −$8,807).

Pooling both connection types yielded a geometric mean ratio of 1.00 (95% CI 0.81–1.25). The pooled arithmetic mean was $136,979 (SD $139,961), compared with $131,837 (SD $129,242), with a difference of +$5,142 (95% CI −$22,362 to +$32,646; P = .71). The pooled state-year standardized difference was +$4,383 (95% CI −$20,531 to +$32,291); additional beneficiary age/risk balancing yielded −$4,900 (95% CI −$29,306 to +$22,584). The pooled intervals included substantial differences in either direction. Contextual state-year standardized annual beneficiary counts were 189.3 versus 186.3, and service units were 595.5 versus 554.7, among all connected and comparison surgeons, respectively.

#### Longitudinal payment trajectories

Among the 50 practice/MSO surgeons eligible for longitudinal analysis, mean payments before and after organizational investment were $238,715 and $187,231, compared with $163,972 and $133,810 among comparison surgeons. The model-based log-payment change was +16.9% (95% CI −0.3% to +37.2%). The dollar change was −$15,727 per year (clustered 95% CI −$78,207 to +$46,753; wild cluster bootstrap 95% CI −$95,893 to +$35,191). Removing the highest preinvestment biller changed the dollar estimate to +$8,556 (95% CI −$13,658 to +$30,770). Parent-omission, upper-tail, and same-ZIP estimates are reported in Figure S1 and Tables S7 and S8. The scale-dependent direction and influence of the upper tail limit interpretation of investment-related change.

Practice/MSO event-study estimates were imprecise. Hospital-operator payments showed a differential change of −$30,848; the coefficient for four or more years before investment was +$43,790 (95% CI +$22,512 to +$65,069), providing evidence inconsistent with parallel trends (Figure 3; Tables S3 and S4).

**Figure 3:**
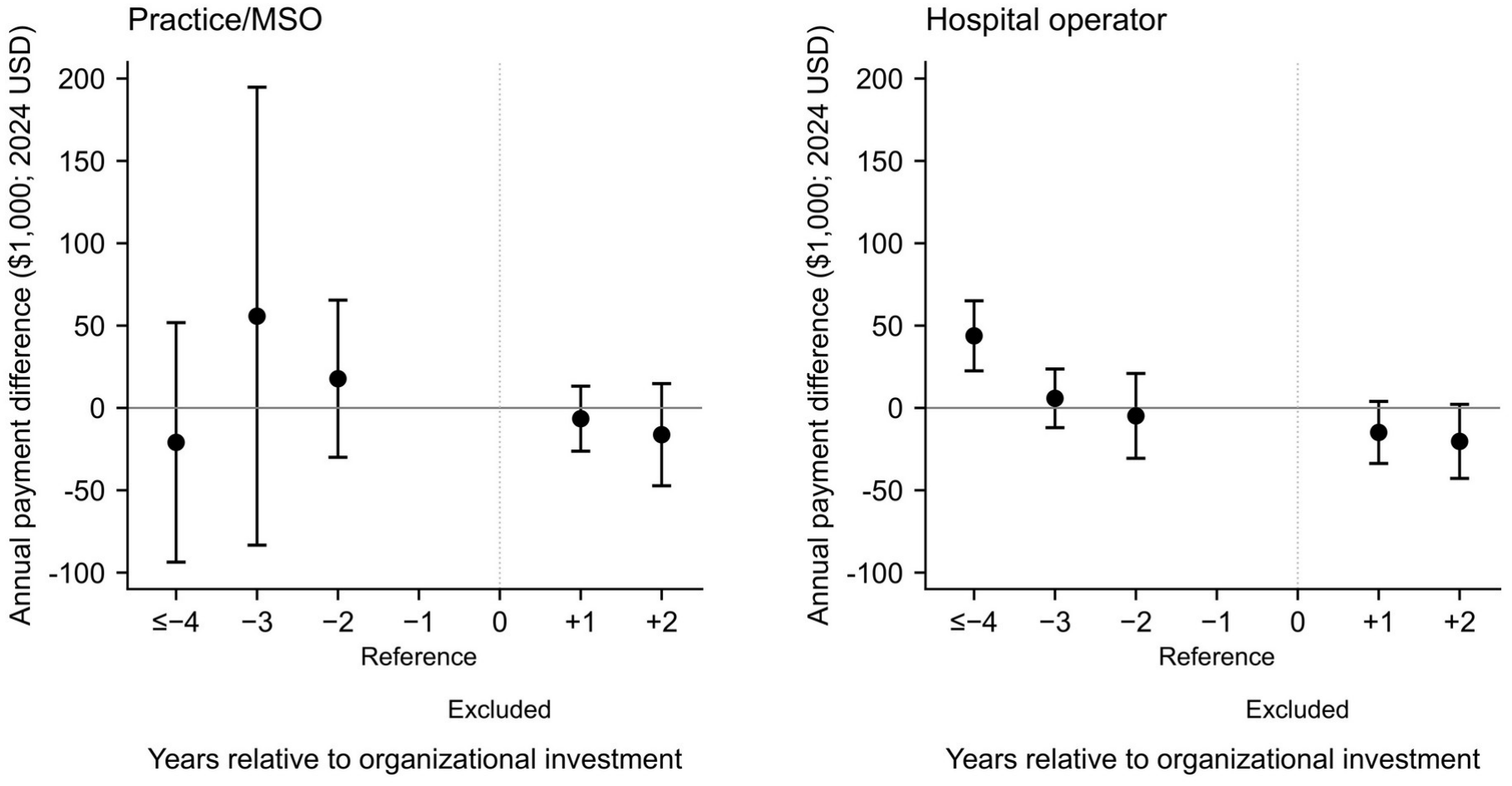
Payment trajectories relative to organizational investment. A, Practice/MSO surgeons (n = 50; 14 connected parent organizations; 2,094 distinct comparison physicians). B, Hospital-operator surgeons (n = 20; 3 connected parent organizations; 1,357 distinct comparison physicians). Points show stacked event-study annual payment differences in 2024 US dollars relative to year −1; bars show clustered-t 95% confidence intervals. Years ≤−4 are pooled; year 0 is excluded. Models use physician-by-subexperiment and year-by-subexperiment fixed effects and comparison sample-share weights. Connected surgeons are clustered by parent organization and comparison physicians by identity across subexperiments; t reference degrees of freedom are 13 and 2. Both panels share the same vertical scale. Hospital-operator preperiod evidence is inconsistent with parallel trends, and inference with three parent organizations is unstable. Practice/MSO preperiod uncertainty does not establish parallel trends. Organizational investment timing does not verify individual employment or acquisition; these trajectories do not establish acquisition effects. MSO, management-services organization. Alt text: Two dollar event-study plots show wide practice/MSO uncertainty and a positive hospital-operator differential four or more years before investment, inconsistent with parallel trends. Year −1 is the reference; investment year 0 is excluded.

## Discussion

Neurosurgeons with documented PE-backed practice or practice-linked MSO connections had higher unadjusted geometric mean Medicare payments than comparison surgeons (ratio 1.38; 95% CI 1.11–1.72), while the age/risk-balanced ratio was 1.25 (95% CI 1.01–1.59), with a lower confidence bound close to unity. The state-year standardized arithmetic mean difference was approximately $32,000. The unstandardized arithmetic mean and median differences were also positive, although their CIs included zero.

### Practice organization and payment profiles

Practice/MSO connections correspond to the physician-practice investment relationships examined in other procedural specialties. Direct acquisitions, minority equity investments, and practice-linked management arrangements can differ in control and operations. The payment difference persisted among metropolitan providers and after omitting influential parent organizations, suggesting that broad rurality or a single large organization did not fully explain the association. Provider-address RUCA codes do not describe patients’ residential geography or the areas served by hospitals.

Hospital-operator surgeons had lower geometric and arithmetic mean payments. Professional-group employment within a hospital system differs from investment in a physician practice or its MSO. Quorum’s fund ownership following restructuring also differs from a conventional practice buyout. Pooling practice/MSO surgeons with lower-payment hospital-operator surgeons reduced the overall contrast. Reporting the groups separately shows how payment levels differ by type of professional connection.

### Relationship to prior specialty studies

Prior studies illustrate why payment levels and acquisition-related changes require separate interpretation. Nie et al. found that urologists later affiliated with PE had higher Medicare payments and patient volume before acquisition.[8] Dongre et al. similarly reported greater procedure volume before investment in otolaryngology, alongside higher payments per procedure after acquisition.[5] The higher preinvestment payment levels among our longitudinally eligible practice/MSO surgeons are consistent with the possibility that investor-linked cohorts differ before organizational investment. They do not establish that selection is the sole explanation, particularly because historical practice membership was not systematically confirmed at the investment date.

Other outcomes also vary across studies. Singh et al. used commercial claims and a matched difference-in-differences design across procedural specialties, finding increases in spending and utilization after acquisition.[11] Braun et al. used Medicare claims in ophthalmology and found little or no overall change in total spending or use, alongside changes in selected clinician and treatment measures.[2] Annual total payment, payment per procedure, and spending per beneficiary capture different dimensions of practice.

### Meaning of annual professional payments

Annual Medicare payments summarize one reimbursed component of a surgeon’s professional activity. Higher payments can reflect a larger beneficiary population or more services per beneficiary. Totals also vary with service mix and payment adjustments. They do not directly measure personal income or practice profit and cannot establish the quality of care. Neurosurgical Medicare research has shown that practice volume and service composition are relevant to payment variation.[9] Here, beneficiary counts were numerically close in the state-year standardized comparison, while service units were higher among connected surgeons. Beneficiary counts and service units help interpret overall payment levels, although the provider summaries do not identify specific procedures or explain why payments differed.

Practice/MSO surgeons had lower mean claims-based risk scores despite higher mean beneficiary age. One hypothesis is that a greater share of their care involves lower-risk candidates for elective procedures, including spine surgery, for which frailty is relevant to perioperative outcomes.[12] CMS risk scores summarize expected spending.[4] They do not measure operative fitness. These provider summaries do not establish surgical indication or urgency, so the elective-care explanation remains untested.

Age/risk balancing attenuated the practice/MSO comparison on both scales. On the same annual-record scale, the geometric mean ratio decreased from 1.31 to 1.25, leaving a point estimate near 25% higher payments. The balanced ratio interval had a lower bound close to, but above, unity; the dollar CI crossed zero. Concentrated comparison weights also limit precision: the largest weight was 10.2 times its base weight, and the annual-record weight concentration measure was 773. Balancing addresses observed provider-level covariate means and state-year composition, while residual differences in procedure mix, referral patterns, and coding may remain.

The longitudinal analysis described payment trajectories around organizational investment among subsequently documented affiliates. Both scales were imprecise, and omitting one high-billing surgeon reversed the dollar estimate. Practice/MSO preperiod uncertainty does not establish parallel trends. The hospital-operator preperiod coefficient provides evidence against that assumption. Same-state and same-ZIP Medicare records do not verify continuous employment at the practice. Historical affiliation dates are needed to interpret these models as acquisition effects on a defined practice cohort.

### Limitations

Public records may miss or misclassify investment connections. Unmeasured differences in subspecialty and practice setting may also influence payments, as may referral patterns. Connected surgeons contributed an index year, whereas comparison surgeons contributed averages over eligible years. Cross-sectional inference and bootstrap intervals assumed independent physicians and did not account for organizational clustering. Age/risk balancing used contemporaneous provider averages and does not provide patient-level adjustment for operative indication or disease severity. The supporting longitudinal analysis used organizational investment timing without systematic confirmation of individual membership at that date and included only 14 practice/MSO and three hospital-operator parent clusters. Medicare fee-for-service payments capture part of practice activity and do not measure compensation or patient outcomes. Multiple comparisons were evaluated without multiplicity adjustment. These descriptive observational comparisons do not establish acquisition effects or payment equivalence.

## Conclusion

Documented PE-backed practice or practice-linked MSO connections were associated with higher unadjusted Medicare professional payment levels among US neurosurgeons. The age/risk-balanced point estimate remained approximately 25% higher, with a lower confidence bound close to unity. Hospital-operator payments were lower. Longitudinal estimates remained imprecise and sensitive to influential observations.

## Data Availability

CMS provider summaries and the CPI-U series are publicly accessible. Supplementary Data 1 provides aggregate analytical results, analysis code, and the organization-level investment bibliography. Physician-level analytical data are available from the corresponding author on request. NPI linkage files, physician-specific source inventories, and original affiliation documents are retained by the authors and are not redistributed.

https://data.cms.gov/provider-summary-by-type-of-service/medicare-physician-other-practitioners

## Abbreviations

CI: confidence interval
CMS: Centers for Medicare & Medicaid Services
MSO: management-services organization
NPI: National Provider Identifier
PE: private equity
SD: standard deviation.

## Disclosures and data availability

### Author contributions

Abhinav Bachu led study design, data collection, analysis, and manuscript writing. Raghav Rajesh contributed to source identification, evidence review, and manuscript writing. Zachary Uram and Anoop Chinthala contributed to clinical interpretation, validation, and critical review and editing of the manuscript. Anand V. Germanwala provided study supervision and manuscript review. All authors approved the final manuscript.

CMS provider summaries and the CPI-U series are publicly accessible.[3,13] Supplementary Data 1 provides aggregate analytical results, analysis code, and the organization-level investment bibliography. Physician-level analytical data are available from the corresponding author on request. NPI linkage files, physician-specific source inventories, and original affiliation documents are retained by the authors and are not redistributed. Study IDs identify individual records discussed in the supplement; the linkage key is not included in shared materials.

## Use of artificial intelligence (AI)-assisted technology for manuscript preparation

During the preparation of this work, the authors used ChatGPT (OpenAI) to assist with language editing and with writing analysis code. AI tools did not independently generate content; all text, code, and analyses were directed, reviewed, edited, and verified by the authors, who take full responsibility for the content of the publication.

