## Supplementary Appendix for "Private Equity Practice Connections and Medicare Professional Payments Among US Neurological Surgeons"

This supplement provides exposure definitions, organization-specific analytical windows, statistical methods, and supporting payment analyses. Table and figure numbers without an S refer to the main manuscript.

**Exposure ascertainment and cohort assembly**

Organizations were identified through PitchBook and targeted searches of public announcements and investment-firm websites through October 2026. One investigator reviewed the evidence for the organizational investment relationship, the surgeon's professional affiliation, and their temporal overlap. Dated practice rosters, archived CMS professional-group records, and professional-affiliation documents were linked by exact individual NPI to the corresponding annual CMS neurosurgery record. Eligibility required professional membership, employment, or a practice-linked economic relationship; hospital privileges or an organization name alone were insufficient.

The source inventory included 10,474 candidate documentary records, not 10,474 unique surgeons. There were 3,046 accepted, 2,130 rejected, and 5,298 unresolved records. Accepted records supported 3,012 eligible dated professional observations after 34 duplicates were removed. Indexing each distinct surgeon at the earliest eligible observation yielded 104 connected surgeons. Unresolved records were omitted from the connected cohort, and known or unresolved professional connections and specified organizational matches excluded corresponding physicians from the comparison pool across years. The search was organization- and roster-based and does not provide a national prevalence estimate.

Table S1 summarizes the parent labels, recorded investors, and eligibility windows represented in the index. These windows define when a dated professional observation could qualify; their endpoints should not be interpreted as universal divestment dates. Multiple starts within a parent label reflect different affiliated organizations. Investment-start dates are distinct from each surgeon's index date and from a confirmed individual acquisition date.

*Classification of selected investment arrangements*

LifePoint qualifies through professional-group employment. The 2018 merger announcement identifies Apollo-affiliated fund ownership (supplemental reference S11). Quorum is explicitly classified as a hospital-operator relationship under investment-fund ownership following its July 2020 restructuring, rather than as an acquired physician practice (S18). BASS qualifies through Privia's majority interest in its exclusive MSO, announced October 18, 2021. Privia's 2022 SEC disclosures document continuing sponsor holdings despite its public listing (S14, S13). All four BASS/Privia index observations are in 2022. Laser Spine Institute qualifies as a practice relationship within the recorded equity window. Its two index observations are in 2015, before its 2019 closure. These classifications concern documented equity and professional links and do not assume uniform investor control or business models.

Every Table S1 row cites investment evidence. The ANS filing documents the 2017 investment (S2); Laser Spine Institute agreements identify equity sponsors (S22). Physician-level affiliation evidence, timing exceptions, and exclusion analyses are documented in the source audit and Table S9.

**Table S1: Organizations and analytical eligibility windows**

| Parent or platform label | Recorded investor | n | Eligibility window |
|---|---|---|---|
| Aligned Orthopedic Partners (S1) | Atlantic Street Capital | 3 | 2022-08-31 to 2022-12-31 |
| ANS Continuum / Altair (S2) | Lorient Capital | 16 | 2017-12-31 to 2022-12-31 |
| BASS Privia MSO / Privia Health (S14, S13) | Goldman Sachs entities; Pamplona | 4 | 2021-10-18 to 2022-12-31 |
| Duly / DMG Practice Management Solutions (S6, S17) | Ares Management | 2 | 2021-12-06 to 2022-12-31 |
| HOPCo (S12, S7) | Audax; Linden; Frazier | 10 | 2021-04-23 to 2023-09-12<br>2022-07-31 to 2023-09-12<br>2022-12-15 to 2023-09-12 |
| iRISE / Florida Spine and Joint Institute (S24) | Trive Capital | 1 | 2018-12-20 to 2021-12-31 |
| Kelsey-Seybold Management Services (S23) | TPG Capital | 3 | 2020-01-21 to 2021-12-31 |
| Laser Spine Institute (S22) | SLG equity vehicles; EFO affiliates | 2 | 2015-07-31 to 2019-03-01 |
| National Spine & Pain Centers (S3) | Avista Capital Partners | 1 | 2017-12-31 to 2021-12-31 |
| OrthoAlliance (S19) | Revelstoke Capital Partners | 8 | 2019-07-23 to 2024-10-31<br>2022-11-16 to 2024-10-31 |
| OrthoNebraska (S9) | InTandem Capital Partners | 1 | 2023-08-02 to 2023-12-31 |
| Orthopedic Care Partners (S27) | Varsity Healthcare Partners | 1 | 2017-11-16 to 2019-11-14 |
| Precision Spine Care / Roseland Management (S4) | Bow River Capital | 2 | 2018-11-09 to 2020-09-30 |
| Prospira PainCare (S15) | Enhanced Equity; Pulse Equity; Webster | 1 | 2013-10-15 to 2020-09-09 |
| Southeastern Spine Institute (S16) | Candescent Partners and co-investors | 1 | 2017-04-19 to 2019-12-31 |
| Spire Orthopedic Partners (S10) | Kohlberg & Company | 5 | 2019-07-08 to 2022-12-31 |
| Summit Health / WP CityMD Topco (S20) | Warburg Pincus; Consonance | 3 | 2019-08-13 to 2022-12-31 |

| | | | |
|---|---|---|---|
| Surgical Affiliates Management Group (S21) | Varsity Healthcare Partners | 1 | 2020-04-14 to 2024-12-31 |
| U.S. Orthopaedic Partners (S25, S8) | FFL Partners | 2 | 2022-05-23 to 2022-12-31 |
| United Musculoskeletal Partners (S26) | Welsh Carson; A&M Capital | 2 | 2022-07-20 to 2023-12-31 |
| LifePoint Health (S11) | Apollo Global Management | 27 | 2018-11-16 to 2024-12-31 |
| Quorum Health / Quincy Health LLC (S18) | Davidson Kempner; GoldenTree | 3 | 2020-07-07 to 2024-12-31 |
| Steward Health Care (S5) | Cerberus Capital Management | 5 | 2010-11-06 to 2020-06-02<br>2017-09-29 to 2020-06-02 |

The first 20 rows are practice/MSO relationships (69 surgeons); the final three hospital-operator rows are LifePoint, Quorum, and Steward (35 surgeons). Dates are the recorded analytical windows, not independently inferred holding periods. References identify organizational investment relationships. Aggregate timing corroboration is summarized in Table S9 and Supplementary Data 1; two index-year assignments remain uncertain. Physician-level affiliation records are retained by the authors.

### Measurement and cross-sectional estimation

The physician is the cross-sectional analysis unit. For a connected surgeon, the payment is the index-year value. For a comparison physician, it is the arithmetic mean of all eligible annual payments. The main contrast exponentiates the difference in mean log payments between practice/MSO and comparison physicians. Supporting dollar contrasts subtract the comparison-physician arithmetic mean from the connected-physician mean. Each physician has equal weight within group. Welch standard errors use the sum of the two sample variances divided by their respective group sizes, with Welch–Satterthwaite degrees of freedom. The practice/MSO log-payment Welch degrees of freedom are 71.6. This permits unequal variance but does not make index-year and multiple-year observation schemes identical.

CMS variables are Tot_Mdcr_Pymt_Amt for payment, Tot_Benes for beneficiaries, Tot_Srvcs for service units, Bene_Avg_Age for beneficiary age, and Bene_Avg_Risk_Scre for risk score. Dollar amounts are multiplied by the ratio of the 2024 annual CPI-U to the annual CPI-U in the observation year before averaging. No diagnosis or HCPCS restriction defines the payment outcome. All cross-sectional payment values are positive, allowing logarithmic analysis. Geometric means exponentiate the means of log physician values; the ratio and CI exponentiate the corresponding Welch log-scale contrast and interval.

Median contrasts subtract group medians and use bootstrap percentile intervals. The two-sided Mann–Whitney test evaluates a distributional/rank contrast; it is not reported as a direct test of the median difference. CIs and P values are nominal and do not use multiplicity corrections. Shared organizational influences are not addressed by these physician-level tests or bootstrap intervals.

*Standardization and bootstrap methods*

For each state-year stratum, the standardized estimator calculates the connected mean minus the mean of comparison annual records, then weights that contrast by the stratum's share of the analyzed connected cohort. The same calculation is repeated using the practice/MSO or hospital-operator stratum distribution for connection-specific estimates. Thus, standardized estimates use annual comparison records and target a different composition from the equal-physician log-payment estimate.

Relative-entropy balancing fits separate comparison weights for the practice/MSO and pooled cohorts. It begins with equal weights within each state-year stratum and stratum totals equal to the analyzed connected distribution, then preserves those totals while matching the cohort's mean beneficiary age and risk score. Payments are not used to fit weights. Practice/MSO targets are age 73.2 years and risk score 1.30; weights cover 5,943 annual comparison records from 3,294 physicians across 36 strata. All weights are positive; the largest is 10.2 times its stratum base weight. The inverse sum of squared normalized record weights is 773. After aggregating weights within physician, the corresponding concentration measure is 575. These summarize weight concentration and are not counts of independent physicians. Pooled-cohort weights match age 72.8 and risk score 1.38, with a maximum weight ratio of 4.8 and concentration measure of 1,783 among 8,118 annual records.

Practice/MSO log contrasts exponentiate the difference between the equal-weighted mean of connected annual log payments and the weighted mean of comparison annual log payments. The state-year-only and age/risk-balanced log estimates use the same aggregation; the main log comparison instead uses each comparison physician's multiyear arithmetic mean before transformation. These are distinct descriptive estimands.

Bootstrap draws independently resample connected and comparison physicians with replacement within group. For the practice/MSO annual-record analyses, the sampling frames comprise 69 connected surgeons and 3,294 comparison physicians contributing 5,943 eligible annual records across 36 state-year strata. This bootstrap conditions on the identified eligible comparison population and available target strata. Every annual record and multiplicity of a sampled comparison physician is retained. Standardization and balancing are refitted in each draw. Percentile 95% CIs use 19,999 draws for practice/MSO-balanced contrasts and the state-year-only log ratio, and 4,999 for other standardized, pooled-balanced, and median estimates. The seed is 20261020; the practice/MSO weighting analysis uses independent PCG64 streams generated by SeedSequence for each draw, sampling connected physicians before comparison physicians. All 19,999 practice/MSO draws had stratum support and converged. Resampling physicians preserves their repeated annual records but not dependence between physicians in a shared organization.

**Table S2: Distributional comparisons of annual payments**

| Group and summary | Payment | Contrast | 95% CI | P |
|---|---|---|---|---|
| Practice/MSO geometric mean | $114,555 | Ratio 1.38 | 1.11 to 1.72 | .004 |
| Practice/MSO median | $117,101 | +$19,007 | −$5,406 to +$70,125 | .030 |
| Hospital operator geometric mean | $43,935 | Ratio 0.53 | 0.35 to 0.81 | .004 |
| Hospital operator median | $58,678 | −$39,416 | −$65,710 to −$25,392 | <.001 |
| All connected geometric mean | $82,975 | Ratio 1.00 | 0.81 to 1.25 | .987 |
| All connected median | $88,690 | −$9,404 | −$25,227 to +$8,210 | .781 |

Comparison geometric mean $82,829 and median $98,094. Ratio CIs and P values use Welch tests on log payments. Median CIs use 4,999 physician-level bootstrap draws; median-row P values are from Mann–Whitney distributional tests. Each connected group is compared with the same full comparison pool.

#### Longitudinal eligibility and model

Longitudinal eligibility requires at least one annual CMS neurosurgery record in the index state before the calendar year of organizational investment and at least one record in the first or second following year. The investment year is excluded. Seventy surgeons meet this rule: 50 practice/MSO and 20 hospital operator. Sixty-five have a preinvestment record in the same ZIP code as their index record, including 46 practice/MSO surgeons. Practice membership at investment start was not systematically confirmed for the longitudinal cohort. Some archived sources describe earlier practice histories, but same-state or same-ZIP Medicare presence alone does not establish employment in the practice at acquisition.

Same-state comparison surgeons must have at least one preinvestment year and at least one year in the two-year postperiod, and must not appear in the comparison-exclusion inventory. A comparison physician can contribute to more than one state–investment-year subexperiment. The practice/MSO stack includes 18 subexperiments, 2,094 distinct comparison physicians, 2,801 comparison memberships, and 20,023 annual observations. The hospital-operator stack includes 12 subexperiments, 1,357 distinct comparison physicians, 1,387 comparison memberships, and 8,789 annual observations. Comparison physician identity is preserved across repeated subexperiments.

The model regresses annual payment on the interaction of connected status with the postperiod, with a separate intercept for each physician within subexperiment and each calendar year within subexperiment. Connected observations receive weight one. Within each subexperiment, comparison observations receive the ratio of that subexperiment's share of all connected physicians to its share of all comparison memberships. The resulting fixed-effect coefficient need not equal a simple subtraction of the displayed group mean changes, particularly with unequal annual follow-up. All available preinvestment years from 2013 are used, with up to two postinvestment years; observation windows are not balanced across physicians. The stacked approach follows the sampling-share weighting framework described by Wing and colleagues.[14]

Cluster-robust covariance uses the cluster-count correction, clustering connected physicians by parent organization and comparison physicians by NPI. The t reference has 13 degrees of freedom for the 14 practice/MSO parent clusters and two degrees of freedom for the three hospital-operator clusters. Clustering comparison physicians preserves their dependence across subexperiments, but not dependence between different physicians in the same comparison practice. Hospital-operator DiD inference is not reported because only three connected parent clusters are available; its postinvestment result is descriptive. Event-study CIs are displayed to assess the preperiod pattern, with the same small-cluster limitation. Event-study models replace the postperiod interaction with indicators for four or more years before, three years before, two years before, one year after, and two years after; the preceding year is the reference. Coefficients describe differences relative to that reference, not unadjusted group means.

**Table S3: Longitudinal comparisons around organizational investment**

| Measure | Practice/MSO | Hospital operator |
|---|---|---|
| Connected surgeons | 50 | 20 |
| Connected parent clusters | 14 | 3 |
| State–investment-year subexperiments | 18 | 12 |

| | | |
|---|---|---|
| Distinct comparison surgeons | 2,094 | 1,357 |
| Connected mean before | $238,715 | $175,909 |
| Connected mean after | $187,231 | $131,129 |
| Comparison mean before | $163,972 | $166,396 |
| Comparison mean after | $133,810 | $152,359 |
| Differential annual dollar change | −$15,727 | −$30,848 |
| Clustered 95% CI | −$78,207 to +$46,753 | Not reported |
| Clustered t P | .596 | Not reported |
| Wild cluster bootstrap 95% CI | −$95,893 to +$35,191 | Not reported |
| Log-scale percentage change | +16.9% | −13.9% |
| Log-scale 95% CI | −0.3% to +37.2% | Not reported |

Annual payments in 2024 US dollars. Models include physician-by-subexperiment and year-by-subexperiment fixed effects and sample-share weights. Group means summarize physician-level pre/post averages and need not algebraically reproduce the regression coefficient. Investment timing refers to the organization, not a verified individual employment/acquisition event. Hospital-operator postinvestment results are point estimates only. Practice/MSO wild cluster bootstrap inference uses 9,999 Webb six-point draws with the null imposed, the same organization/physician clusters, and both fixed effects re-estimated in each draw; the CI inverts the two-sided bootstrap t test. Other practice/MSO CIs use the clustered t procedure.

**Table S4: Event-study estimates relative to the year before investment**

| Event time | Practice/MSO estimate and 95% CI | Hospital-operator estimate and 95% CI |
|---|---|---|
| Four or more years before | −$20,921<br>(−$93,645 to +$51,804) | +$43,790<br>(+$22,512 to +$65,069) |
| Three years before | +$55,717<br>(−$83,329 to +$194,764) | +$5,832<br>(−$11,982 to +$23,645) |
| Two years before | +$17,749<br>(−$29,972 to +$65,471) | −$4,823<br>(−$30,593 to +$20,947) |
| One year after | −$6,519<br>(−$26,263 to +$13,224) | −$14,875<br>(−$33,717 to +$3,967) |
| Two years after | −$16,257<br>(−$47,282 to +$14,769) | −$20,310<br>(−$42,796 to +$2,176) |

Investment year excluded. Estimates are annual payment differences in 2024 US dollars relative to the year before organizational investment. CIs use the clustered t procedure described above. Practice/MSO preperiod estimates are imprecise and do not confirm parallel trends. For hospital operators, the coefficient four or more years before investment is positive with a CI excluding zero, providing evidence inconsistent with parallel trends. Hospital-operator CIs use only three connected parent clusters and are unstable.

**Geographical sensitivity**

CMS Rndrng_Prvdr_RUCA was retrieved from the original annual by-provider datasets and joined by NPI and year. Matching state and inflation-adjusted payment values were checked against the analytical records. Primary RUCA codes 1–3 define metropolitan provider addresses and 4–10 nonmetropolitan addresses. All 104 connected records and 8,117 of 8,118 comparison annual records had usable codes; the one missing record was the sole observation for one comparison physician. Within each geography, comparison physicians contribute their mean over eligible records in that geography, whereas connected surgeons contribute their index record. A comparison physician can appear in both strata. This measures provider-address geography rather than patient residence or hospital catchment area.

**Table S5: Payment comparisons stratified by provider RUCA**

| Provider geography and group | Group n | Control n | Mean difference and 95% CI | Geometric mean ratio and 95% CI |
|---|---|---|---|---|
| Metropolitan<br>All connected | 98 | 3956 | +$131<br>(−$26,458 to +$26,720) | 0.99<br>(0.80 to 1.23) |
| Metropolitan<br>Practice/MSO | 68 | 3956 | +$33,673<br>(−$826 to +$68,173) | 1.42<br>(1.15 to 1.76) |
| Metropolitan<br>Hospital operator | 30 | 3956 | −$75,897<br>(−$95,247 to −$56,547) | 0.44<br>(0.29 to 0.65) |
| Nonmetropolitan<br>All connected | 6 | 71 | +$75,078<br>(−$182,310 to +$332,467) | 1.08<br>(0.21 to 5.58) |
| Nonmetropolitan<br>Practice/MSO | 1 | 71 | −$130,499<br>(CI not estimated) | Not estimated |
| Nonmetropolitan<br>Hospital operator | 5 | 71 | +$116,194<br>(−$193,425 to +$425,813) | 1.53<br>(0.25 to 9.36) |

Payments are 2024 US dollars. Welch intervals use physician-level values; ratio intervals use log payments. The nonmetropolitan practice/MSO stratum has one surgeon, precluding variance estimation. Its mean is $17,960, compared with $148,459 among nonmetropolitan comparison physicians. Only six connected surgeons are nonmetropolitan, so subgroup results are particularly uncertain.

**Influence and continuity analyses**

Targeted cross-sectional omissions remove ANS Continuum/Altair from the practice/MSO group and LifePoint from the pooled or hospital-operator groups while retaining the original comparison pool. The longitudinal analysis omits each of the 14 practice/MSO parents in turn, rebuilding subexperiments and comparison sample-share weights. Upper-tail analyses rank the 50 eligible practice/MSO surgeons by their individual mean preinvestment payment, excluding values above the empirical 99th or 95th percentile (one or three surgeons, respectively). These surgeons account for 11.4% or 24.9% of the sum of the 50 preinvestment physician means. Trimming the treated cohort alone changes the target population and is an influence diagnostic, not a corrected causal estimate. The same-ZIP restriction requires at least one preinvestment CMS record with the index ZIP; it does not confirm continuous employment at the practice.

**Table S6: Cross-sectional omission of influential parent organizations**

| Connected group | n | Mean difference and 95% CI | Geometric mean ratio and 95% CI |
|---|---|---|---|
| Practice/MSO without ANS | 53 | +$43,258<br>(+$781 to +$85,734) | 1.47<br>(1.14 to 1.89) |
| All connected without LifePoint | 77 | +$20,486<br>(−$11,167 to +$52,138) | 1.26<br>(1.02 to 1.56) |
| Hospital operators without LifePoint | 8 | −$72,358<br>(−$109,344 to −$35,373) | 0.56<br>(0.30 to 1.06) |

Each group is compared with the original 4,016 comparison physicians. Intervals use Welch procedures on dollars or log payments. These omissions assess dependence on cohort composition; cross-sectional intervals do not account for clustering within organizations.

**Table S7: Practice and MSO longitudinal sensitivity analyses**

| Specification | n | Parent n | Annual dollar change and 95% CI | Log-scale change and 95% CI |
|---|---|---|---|---|
| Full cohort | 50 | 14 | −$15,727<br>(−$78,207 to +$46,753) | +16.9%<br>(−0.3% to +37.2%) |
| Above 99th percentile removed | 49 | 14 | +$8,556<br>(−$13,658 to +$30,770) | +20.6%<br>(+4.2% to +39.5%) |
| Above 95th percentile removed | 47 | 13 | +$21,277<br>(+$181 to +$42,373) | +23.6%<br>(+5.8% to +44.3%) |
| Same-ZIP preinvestment record | 46 | 14 | −$19,375<br>(−$86,729 to +$47,979) | +14.7%<br>(−2.4% to +34.9%) |

CIs in this table use the clustered t procedure. The additional full-cohort wild cluster bootstrap CI is −$95,893 to +$35,191. Controls and weights are reassembled after each restriction. Above-99th-percentile removal retains 18 subexperiments and 2,094 distinct comparison physicians. Above-95th-percentile removal retains 17 subexperiments and 1,813 distinct comparison physicians. The same-ZIP restriction retains 17 subexperiments and 1,848 distinct comparison physicians. The trimmed estimates assess influence and do not supersede the full-cohort dollar or log models.

**Table S8: Practice and MSO leave one parent out analysis**

| Parent omitted | n kept | Annual dollar change | 95% CI |
|---|---|---|---|
| ANS Continuum / Altair | 38 | −$20,794 | −$101,824 to +$60,236 |
| Aligned Orthopedic Partners | 47 | −$20,565 | −$88,317 to +$47,187 |
| BASS Privia MSO / Privia Health | 49 | −$15,686 | −$79,107 to +$47,736 |
| HOPCo | 42 | −$19,491 | −$96,591 to +$57,609 |
| Kelsey-Seybold Management Services | 47 | −$19,042 | −$86,423 to +$48,339 |
| Laser Spine Institute | 48 | −$15,885 | −$80,777 to +$49,007 |
| OrthoAlliance | 43 | −$20,407 | −$93,889 to +$53,076 |
| Orthopedic Care Partners | 49 | −$10,685 | −$73,137 to +$51,767 |
| Precision Spine Care / Roseland Management | 48 | +$11,622 | −$10,214 to +$33,457 |
| Southeastern Spine Institute | 49 | −$20,047 | −$84,409 to +$44,314 |
| Spire Orthopedic Partners | 45 | −$24,513 | −$93,899 to +$44,873 |
| Surgical Affiliates Management Group | 49 | −$16,758 | −$81,103 to +$47,586 |
| U.S. Orthopaedic Partners | 48 | −$14,805 | −$80,638 to +$51,027 |
| United Musculoskeletal Partners | 48 | −$15,903 | −$81,355 to +$49,549 |

Each model is rebuilt after omitting one connected parent. CIs use the clustered t procedure with the retained number of connected parents minus one degrees of freedom. ANS Continuum/Altair accounts for 12 of the 50 longitudinally eligible surgeons and 16 of the 69 cross-sectional practice/MSO surgeons. Omitting Precision Spine Care/Roseland changes the point estimate to +$11,622; the overall range is −$24,513 to +$11,622.

**Source documentation and timing sensitivity**

A physician-level source audit corroborated 81 index-year affiliations using dated CMS Doctors & Clinicians snapshot rows with matching organization identifiers. Of the remaining 23 surgeons, 17 had a retained dated organization or professional record within the recorded investment window and five had corroborating in-window CMS affiliations. Thus, 103 surgeons had affiliation corroboration within the organizational investment window.

Aggregate audit results are provided in Supplementary Data 1. Physician-specific source inventories and affiliation documents are retained by the authors and are not redistributed.

Calendar-year review supported 102 of the 104 assigned index years using dated records or explicit historical employment information. Study ID P103 had a dated professional history supporting the index year despite later roster captures. The earliest retained roster for study ID P074 was from the following calendar year, 33 days after the recorded investment window ended. The retained roster for study ID P080 was within the investment window but from the calendar year after the assigned index year. Self-reported start dates were not treated as independent corroboration of these two index years. Neither P074 nor P080 was longitudinally eligible.

Source-timing exclusions omitted P074 alone or both P074 and P080 from the full 104-surgeon cohort (Table S9). Both retained the original comparison pool and its exclusion rules. Neither changed the longitudinal cohorts or estimates. These exclusions assess the payment comparison without assuming that a later roster establishes an earlier affiliation.

**Table S9: Cross-sectional comparisons after source timing exclusions**

| Exclusion and group | n | Mean difference and 95% CI | Geometric mean ratio and 95% CI |
|---|---|---|---|
| Omit P074<br>Practice/MSO | 68 | +$33,384<br>(−$1,112 to +$67,881) | 1.42<br>(1.15 to 1.76) |
| Omit P074<br>All connected | 103 | +$6,297<br>(−$21,376 to +$33,971) | 1.02<br>(0.82 to 1.26) |
| Omit P074 and P080<br>Practice/MSO | 67 | +$29,645<br>(−$4,543 to +$63,833) | 1.39<br>(1.13 to 1.73) |
| Omit P074 and P080<br>All connected | 102 | +$3,576<br>(−$23,831 to +$30,982) | 1.00<br>(0.81 to 1.24) |

Each group is compared with the original 4,016 comparison physicians. Dollar and log-payment CIs use Welch procedures and do not account for shared organizations. The 35-surgeon hospital-operator group is unchanged in both exclusions. Neither omitted surgeon enters a longitudinal model.

**Figure S1: Practice/MSO longitudinal influence and continuity analyses**

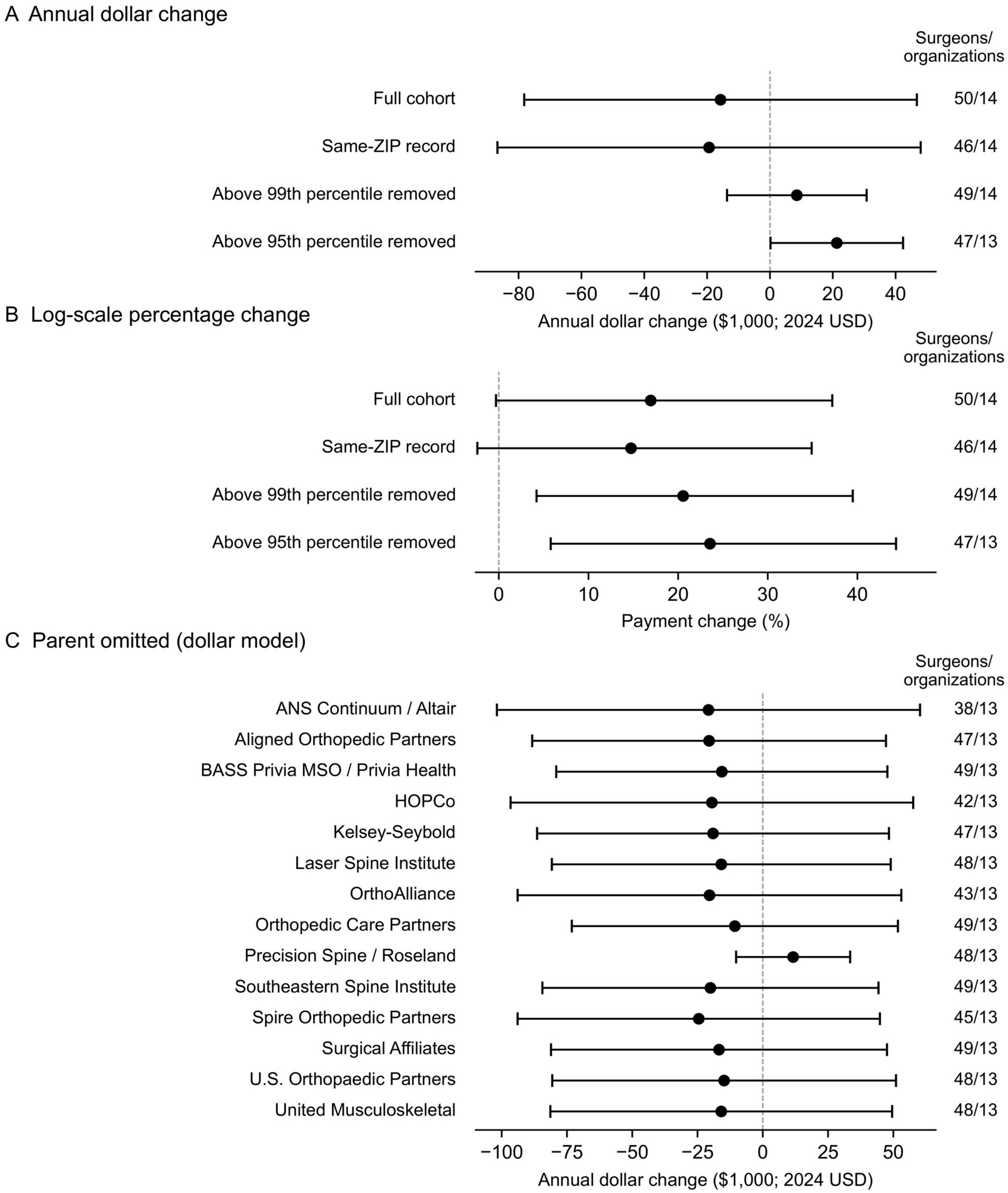


A and B, Full-cohort, same-ZIP, and upper-tail exclusion specifications use state-investment-year stacks, sample-share weights, physician-by-subexperiment and year-by-subexperiment fixed effects, and parent/comparison-physician clustering. Bars show clustered-t 95% confidence intervals, with retained connected organizations minus one degrees of freedom. Panel B displays percentage changes derived from a log-payment model as 100 × [exp(coefficient) − 1]; both CI endpoints receive the same transformation and the displayed percentage axis is linear. C, Each connected parent organization is omitted in turn. Retained surgeon and organization counts are shown. Dollar estimates are in 2024 US dollars. The full-cohort restricted wild cluster bootstrap 95% CI is −$95,893 to +$35,191 (9,999 Webb draws, null imposed, both fixed effects refitted); it is separate from the clustered-t intervals plotted. Upper-tail removal uses preinvestment mean payments and changes the target population; it is an influence diagnostic, not a corrected causal estimate. Same-ZIP records do not verify continuous employment. MSO, management-services organization; ZIP, postal ZIP code.

Alt text: Separate dollar and percentage-change sensitivity panels examine same-ZIP restrictions and upper-tail removals, with a dollar leave-one-organization-out panel. Removing influential high-billing surgeons reverses the full-cohort dollar point estimate. Percentage changes are derived from log models and plotted on a linear axis.

**Supplemental source references**

S1. Aligned Orthopedic Partners. Aligned Orthopedic Partners announces strategic partnership with Jordan-Young Institute. August 18, 2022. Available from: https://www.prnewswire.com/news-releases/aligned-orthopedic-partners-announces-strategic-partnership-with-jordan-young-institute-301608474.html

S2. Atlantic Neurosurgical Specialists, P.A., et al. Combined joint disclosure statement and joint Chapter 11 plan of liquidation. US Bankruptcy Court, District of New Jersey; case 24-15726-VFP, document 223. October 2, 2024:4. Describes the 2017 Lorient investment and October 11, 2017 management agreement. Available from: https://elevenflo.com/documents/3d29566e-9bdb-4c5d-b89b-a8fd7e943a07/

S3. Avista Capital Partners. Avista Capital Partners has acquired National Spine & Pain Centers. June 5, 2017. Available from: https://www.avistahealthcare.com/news_item/avista-capital-partners-has-acquired-national-spine-pain-centers/

S4. Bow River Capital. Bow River Capital completes recapitalization of Roseland Management and Precision Spine Care. November 14, 2018. Available from: https://www.globenewswire.com/news-release/2018/11/14/1651895/0/en/bow-river-capital-completes-recapitalization-of-roseland-management-and-precision-spine-care.html

S5. Cerberus Capital Management. Letter to the Senate Committee on Health, Education, Labor, and Pensions regarding Steward Health Care. September 2024. Describes investment history and exit in 2020. Available from: https://www.cerberus.com/media/2024/09/Letter-to-Senate-HELP-Committee.pdf

S6. DuPage Medical Group. DuPage Medical Group announces investment from Ares Management Private Equity Group. August 16, 2017. Available from: https://www.dulyhealthandcare.com/news/dupage-medical-group-announces-investment-from-ares-management-private-equity-group

S7. Edgemont Partners. Edgemont Partners advises Spine Nevada on partnership with HOPCo. May 6, 2021. Reports April 23, 2021 transaction closing. Available from: https://www.edgemont.com/news/edgemont-partners-advises-spine-nevada-on-partnership-with-hopco/

S8. FFL Partners. U.S. Orthopaedic Partners announces two new acquisitions. March 2, 2021. Describes FFL's October 2020 platform investment. Available from: https://www.fflpartners.com/u-s-orthopaedic-partners-announces-two-new-acquisitions/

S9. InTandem Capital Partners. InTandem Capital Partners announces strategic investment in OrthoNebraska. August 2, 2023. Available from: https://intandemcapital.com/news-and-videos/intandem-capital-partners-announces-strategic-investment-in-orthonebraska/

S10. Kohlberg & Company. ONS announces growth capital partnership with Kohlberg & Company. July 8, 2019. Available from: https://www.kohlberg.com/ons-announces-growth-capital-partnership-with-kohlberg-company/

S11. LifePoint Health. LifePoint Health and RCCH HealthCare Partners announce completion of merger. November 16, 2018. SEC exhibit 99.1. Available from: https://www.sec.gov/Archives/edgar/data/1301611/000119312518329058/d654482dex991.htm

S12. Linden Capital Partners. Healthcare Outcomes Performance Company. Portfolio record; investment since August 2019. Available from: https://www.linden.com/portfolio/investments/current/hopco/

S13. Privia Health Group, Inc. Annual report for the year ended December 31, 2022. Form 10-K. Filed 2023. Available from: https://www.sec.gov/Archives/edgar/data/1759655/000175965523000026/prva-20221231.htm

S14. Privia Health. Privia Health announces entry into California and West Texas markets. October 18, 2021. Available from: https://ir.priviahealth.com/news-releases/news-release-details/privia-health-announces-entry-california-and-west-texas-markets/

S15. Prospira PainCare. Prospira PainCare expands pain management services into Midwestern region. October 15, 2013. Identifies Neuro Pain Consultants and the platform's equity sponsors. Available from: https://www.prnewswire.com/news-releases/prospira-paincare-expands-pain-management-services-into-midwestern-region-227805791.html

S16. Provident Healthcare Partners. Provident Healthcare Partners advises Southeastern Spine Institute on its recapitalization by Candescent Partners. April 19, 2017. Available from: https://www.prnewswire.com/news-releases/provident-healthcare-partners-advises-southeastern-spine-institute-on-its-recapitalization-by-candescent-partners-300441525.html

S17. Quincy Medical Group. Healthcare transformation continues as Quincy Medical Group–Duly Health and Care partnership becomes official. December 6, 2021. Available from: https://www.quincymedgroup.com/news/healthcare-transformation-continues-as-quincy-medical-group-duly-health-and-care-partnership-becomes-official

S18. Quorum Health Corporation. Quorum Health successfully completes financial restructuring. July 7, 2020. SEC exhibit 99.2. Available from: https://www.sec.gov/Archives/edgar/data/1650445/000119312520188872/d942866dex992.htm

S19. Revelstoke Capital Partners. Revelstoke and Beacon Orthopaedics launch national management services organization. July 23, 2019. Available from: https://revelstokecapital.com/news/revelstoke-and-beacon-orthopaedics-launch-national-management-services-organization/

S20. Summit Health. CityMD and Summit Health finalize merger. August 13, 2019. Available from: https://www.summithealth.com/news/citymd-and-summit-health-finalize-merger

S21. Summit Partners. Summit exits Surgical Affiliates Management Group. April 14, 2020. Available from: https://www.summitpartners.com/news/summit-exits-surgical-affiliates-management-group

S22. Texas Capital Bank, N.A. Proof of claim, Laser Spine Institute, LLC; attached credit agreements. 2019. Sponsor Group definitions identify EFO Laser Spine Institute, Ltd. and SLG LSI Investment LLC (PDF pp 102 and 361). Available from: https://lsi-assignee.com/wp-content/uploads/2019/06/POC-Laser-Spine-Institute-LLC.pdf

S23. TPG. Kelsey-Seybold Clinic partners with TPG with plans for expansion of accountable care model in Houston. January 21, 2020. Available from: https://www.tpg.com/news-and-insights/kelsey-seybold-clinic-partners-tpg-plans-expansion-accountable

S24. Trive Capital. Trive Capital partners with Florida Spine and Joint Institute. December 2018. Available from: https://www.trivecapital.com/trive-capital-partners-with-florida-spine-and-joint-institute/

S25. U.S. Orthopaedic Partners. U.S. Orthopaedic Partners expands network in Alabama, adds SportsMED Orthopedic Surgery & Spine Center. May 23, 2022. Available from: https://www.prnewswire.com/news-releases/us-orthopaedic-partners-expands-network-in-alabama-adds-sportsmed-orthopedic-surgery--spine-center-301552462.html

S26. United Musculoskeletal Partners. United Musculoskeletal Partners merge with Novum Orthopedic Partners and addition of founding practice. July 20, 2022. Business Wire release reproduced by citybiz. Available from: https://www.citybiz.co/article/296058/united-musculoskeletal-partners-merge-with-novum-orthopedic-partners-and-addition-of-founding-practice/

S27. Varsity Healthcare Partners. Orthopedic Care Partners. Portfolio record; investment year 2017. Available from: https://varsityhealthcarepartners.com/portfolio/orthopedic-care-partners/

Supplemental web sources accessed October 19, 2026. Main references 3, 13, and 14 provide CMS definitions, CPI-U, and the stacked-analysis framework. Organization-level sources substantiate investment relationships. Aggregate timing-audit results are provided in Supplementary Data 1; the two index-year exceptions are addressed using study IDs in Table S9. Physician-specific affiliation sources are retained by the authors.
