## Supplementary Data 1 (PDF) for "Private Equity Practice Connections and Medicare Professional Payments Among US Neurological Surgeons"

Private Equity Practice Connections and Medicare Professional Payments Among US Neurological Surgeons

Tabular CSV contents are provided in Supplementary_Data_1.xlsx (one sheet per table, plus bibliography sheets).

#### 1. README

SUPPLEMENTARY DATA 1
Private Equity Practice Connections and Medicare Professional Payments
Among US Neurological Surgeons
Release assembled November 9, 2026

CONTENTS
aggregate_results/
Tables 1-3 and S1-S9 as CSV, with table context.
Weighted-analysis and longitudinal aggregate estimates as JSON.
timing_audit/
Aggregate corroboration counts and explanation of the two timing exceptions.
investment_bibliography/
The 23 organization/platform rows and 27 references with original source URLs.
code/
Analytical algorithms, a runnable driver, input schema, synthetic smoke test,
requirements, and source-code provenance.
VALIDATION.json / MANIFEST.json
Packaging checks and SHA-256 checksums.

WHAT WAS VERIFIED
All twelve tables were exported from the current manuscript. All 27
bibliography URLs were extracted from its original hyperlinks.
The three updated weighted contrasts and the corrected practice/MSO
wild-bootstrap interval match saved analytical outputs at manuscript precision.
The algorithms pass synthetic smoke tests. The ZIP passes an integrity test.
This is not a new independent full reanalysis or fresh source adjudication.
Some table results are manuscript transcriptions, identified as such.

DATA RESTRICTIONS
No physician-level payment rows, NPIs, physician names, linkage key, physician
source inventories, affiliation documents, source-archive downloads, or
physician bootstrap draws are included. Small organization-level counts and
the manuscript's public study IDs in Table S9 are included intentionally.
Physician-level analytical inputs must be requested from the corresponding
author subject to the manuscript's data-availability conditions.
This package does not claim that proprietary investment-database records can
be redistributed or that CMS alone reconstructs the curated exposure cohort.

HOW TO USE
1. Review CSV/JSON files without installing software.
2. To test the code:
python -m venv .venv
### Activate the environment using your operating system's command.
python -m pip install -r code/requirements.txt
python code/smoke_test.py
3. To rerun analyses after obtaining the required private analytical inputs:
python code/run_analysis.py --input-dir /path/to/inputs --output-dir /path/to/results
This runs Welch comparisons, median/rank summaries, state-year dollar point
estimates, and the practice/MSO weighting bootstrap (19,999 draws).
Add --longitudinal to run stacked models, event studies, upper-tail and

same-ZIP restrictions, parent omissions, and the corrected 9,999-draw practice/MSO wild-bootstrap inversion.
Optional --draws 99 is for debugging only, NOT publication inference.
An unsupported/bootstrap convergence failure stops the run; no redraws.

REPRODUCIBILITY BOUNDARIES

The published weighted intervals reflect an explicitly specified rebuilt conditional analysis with 3,294 eligible comparison physicians and independent PCG64 streams per draw. Historical bootstrap code/RNG order was not recovered.
The package does not claim exact replication of superseded manuscript intervals.
The source-documentation counts and Tables S5, S6, and S9 are supplied as aggregate manuscript results. Their original source-ascertainment/geography/timing workflows are not reconstructed by the generic driver; full independent reproduction requires retained sources, RUCA joins, and timing adjudication.
The original pooled/standardized/median 4,999-draw intervals are supplied in tables; the driver does not claim to recover their exact historical RNG stream.
Table S3 group means are supplied as reported; fixed-effect coefficients need not equal a simple difference between displayed group mean changes.

METHOD INTERPRETATION

Welch and cross-sectional bootstrap inference assumes independent physicians and does not adjust for shared organizations. Weighted annual-record contrasts and equal-physician multiyear summaries are distinct estimands.
Longitudinal dates mark organizational investment, not verified individual employment/acquisition. Practice/MSO preperiod uncertainty does not establish parallel trends; hospital-operator preperiod evidence is inconsistent with parallel trends. Hospital-operator postperiod results are point estimates only because there are three connected parent organizations.
Analyses are descriptive and do not establish acquisition effects.

RELATIONSHIP TO THE MANUSCRIPT

This package accompanies, rather than replaces, the manuscript's Supplementary Methods and Results and figure artwork.

#### 2. Table Context

Tables transcribed directly from the current manuscript export; they are not newly estimated.
CSV cells may contain line breaks. All dollar outcomes are 2024 USD.

Table 1: Cohort and beneficiary characteristics
The main exposure group comprises practice/MSO surgeons. Hospital-operator surgeons are evaluated separately; their union forms the 104-surgeon pooled cohort. State and state-year counts overlap between connection types. Comparison surgeons are drawn from the 52 state-year strata represented by the full documented-connection inventory.

Table 2: Physician-level annual Medicare professional payment comparisons
Dollar contrasts are connected-minus-comparison arithmetic mean differences in 2024 US dollars. Log contrasts are geometric mean ratios. Comparison arithmetic and geometric means are $131,837 and $82,829 among 4,016 physicians. Welch intervals allow unequal variances and assume independent physicians. Each connected surgeon contributes an index year; each comparison surgeon contributes the arithmetic mean of eligible years before log transformation. P values are nominal and unadjusted for multiple comparisons.

Table 3: Standardized and beneficiary-balanced payment comparisons
Dollar contrasts are differences in arithmetic mean annual payments in 2024 US dollars. Ratios compare geometric means. Standardization uses each connected group's state-year distribution; age/risk balance additionally matches that group's mean provider-level beneficiary age and risk score. Weighted comparisons use annual comparison records. Their log ratios therefore differ in aggregation and weighting from the main physician-summary ratio in Table 2.

Table S1: Organizations and analytical eligibility windows
The first 20 rows are practice/MSO relationships (69 surgeons); the final three hospital-operator rows are LifePoint, Quorum, and Steward (35 surgeons). Dates are the recorded analytical windows, not independently inferred holding periods. References identify organizational investment relationships. Aggregate timing corroboration is summarized in Table S9 and Supplementary Data 1; two index-year assignments remain uncertain. Physician-level affiliation records are retained by the authors.

Table S2: Distributional comparisons of annual payments
Comparison geometric mean $82,829 and median $98,094. Ratio CIs and P values use Welch tests on log payments. Median CIs use 4,999 physician-level bootstrap draws; median-row P values are from Mann–Whitney distributional tests. Each connected group is compared with the same full comparison pool.

Table S3: Longitudinal comparisons around organizational investment
Annual payments in 2024 US dollars. Models include physician-by-subexperiment and year-by-subexperiment fixed effects and sample-share weights. Group means summarize physician-level pre/post averages and need not algebraically reproduce the regression coefficient. Investment timing refers to the organization, not a verified individual employment/acquisition event. Hospital-operator postinvestment results are point estimates only. Practice/MSO wild cluster bootstrap inference uses 9,999 Webb six-point draws with the null imposed, the same organization/physician clusters, and both fixed effects re-estimated in each draw; the CI inverts the two-sided bootstrap t test. Other practice/MSO CIs use the clustered t procedure.

Table S4: Event-study estimates relative to the year before investment
Investment year excluded. Estimates are annual payment differences in 2024 US dollars relative to the year before organizational investment. CIs use the clustered t procedure described above. Practice/MSO preperiod estimates are imprecise and do not confirm parallel trends. For hospital operators, the coefficient four or more years before investment is positive with a CI excluding zero, providing evidence inconsistent with parallel trends. Hospital-operator CIs use only three connected parent clusters and are unstable.

Table S5: Payment comparisons stratified by provider RUCA
Payments are 2024 US dollars. Welch intervals use physician-level values; ratio intervals use log payments. The nonmetropolitan practice/MSO stratum has one surgeon, precluding variance estimation. Its mean is $17,960, compared with $148,459 among nonmetropolitan comparison physicians. Only six connected surgeons are nonmetropolitan, so subgroup results are particularly uncertain.

Table S6: Cross-sectional omission of influential parent organizations
Each group is compared with the original 4,016 comparison physicians. Intervals use Welch procedures on dollars or log payments. These omissions assess dependence on cohort composition; cross-sectional intervals do not account for clustering within organizations.

Table S7: Practice and MSO longitudinal sensitivity analyses
CIs in this table use the clustered t procedure. The additional full-cohort wild cluster bootstrap CI is –$95,893 to +$35,191.

Controls and weights are reassembled after each restriction. Above-99th-percentile removal retains 18 subexperiments and 2,094 distinct comparison physicians. Above-95th-percentile removal retains 17 subexperiments and 1,813 distinct comparison physicians. The same-ZIP restriction retains 17 subexperiments and 1,848 distinct comparison physicians. The trimmed estimates assess influence and do not supersede the full-cohort dollar or log models.

Table S8: Practice and MSO leave one parent out analysis

Each model is rebuilt after omitting one connected parent. CIs use the clustered t procedure with the retained number of connected parents minus one degrees of freedom. ANS Continuum/Altair accounts for 12 of the 50 longitudinally eligible surgeons and 16 of the 69 cross-sectional practice/MSO surgeons. Omitting Precision Spine Care/Roseland changes the point estimate to +$11,622; the overall range is –$24,513 to +$11,622.

Table S9: Cross-sectional comparisons after source timing exclusions

Each group is compared with the original 4,016 comparison physicians. Dollar and log-payment CIs use Welch procedures and do not account for shared organizations. The 35-surgeon hospital-operator group is unchanged in both exclusions. Neither omitted surgeon enters a longitudinal model.

Note: Full CSV tables (Table_1–3, Table_S1–S9) and bibliography CSVs (sources, organization_windows) are in Supplementary_Data_1.xlsx.

### 3. Aggregate JSON Results

#### *weighted_analysis.json*


```
{
  "origin": "Saved rebuilt analysis, explicitly adopted in the current manuscript",
  "historical_replication": "Original 19999-draw bootstrap sampling frame and RNG order were not recovered; this is an explicit conditional ana[illegible]
  "population": {
    "connected_physicians": 69,
    "comparison_physicians": 3294,
    "comparison_annual_records": 5943,
    "state_year_strata": 36
  },
  "estimates": {
    "state_year_ratio": {
      "estimate": 1.3055162585874358,
      "ci": [
        1.0666209818985328,
        1.5972086432585146
      ]
    },
    "balanced_ratio": {
      "estimate": 1.254073661083553,
      "ci": [
        1.0063220131612975,
        1.588734718874501
      ]
    },
    "balanced_dollar_difference": {
      "estimate": 19707.652474939736,
      "ci": [
        -10418.633782219653,
        52918.55971049494
      ]
    }
  },
  "balance_diagnostics": {
    "max_standardized_balance_error": 4.1648153520544474e-14,
    "max_stratum_mass_error": 2.7755575615628914e-17,
    "iterations": 6,
    "fallback": false,
    "target_means": [
      73.21739130434783,
      1.3044275362318838
    ],
    "weighted_means": [
      73.21739130434796,
      1.3044275362318856
    ]
  },
  "weight_concentration": {
    "state_year_only": {
      "minimum_record_weight": 3.3316674995835414e-05,
      "maximum_record_weight": 0.0019698888419867736,
      "maximum_ratio_to_state_year_weight": 1.0,
      "record_effective_n": 1425.0372014222635,
      "physician_effective_n": 1019.3790591267417,
      "maximum_physician_weight": 0.0024276572696009238,
      "top_1_percent_record_mass": 0.11819333051920641,
      "top_10_record_mass": 0.01969888841986774
    },
    "age_risk_balanced": {
      "minimum_record_weight": 3.1877616740358526e-09,
      "maximum_record_weight": 0.009912890808538863,
      "maximum_ratio_to_state_year_weight": 10.20147139220453,
      "record_effective_n": 773.2745457605048,
      "physician_effective_n": 574.8439993544403,
      "maximum_physician_weight": 0.010453747048189714,
      "top_1_percent_record_mass": 0.2092100592736575,
      "top_10_record_mass": 0.06477971206862061
    }
  },
  "bootstrap": {
    "requested_draws": 19999,
    "successful_draws": 19999,
    "seed": 20261020,
    "interval": "95% percentile, numpy linear quantile (type 7)",
    "unit": "physician, independently within connected/comparison groups",
    "maximum_standardized_balance_error": 9.980085646787984e-10,
    "maximum_stratum_mass_error": 3.3306690738754696e-16
  },
  "specification": {
    "version": "2026-10-28-method-defined-review",
```

"analysis": "Practice/MSO annual-record payment contrasts",
"decision_basis": "Define the analytical population and conditioning before interpreting CI endpoints; do not choose a bootstrap frame by a
"target": "Descriptive contrast between 69 connected index-year records and annual comparison log payments standardized to the connected co
"comparison_population": {
"eligible_physicians": 3294,
"eligible_annual_records": 5943,
"state_year_strata": 36
},
"selected_bootstrap_frame": "3294 comparison physicians with at least one record eligible for this practice/MSO estimand",
"conditioning": "Condition on the identified empirical practice/MSO comparison population and its size, and on the set of available target
"justification": "Physicians with no record in the 36 target strata cannot contribute to this estimator. Resampling the eligible empirical
"full_pool_alternative": "Resampling 4016 then retaining eligible annual records randomly varies the number of eligible sampled physicians.
"not_estimated": [
"Uncertainty in source ascertainment or investment classification",
"Uncertainty from discovering the analytical target strata",
"Dependence between physicians sharing organizations",
"Causal acquisition effects"
],
"bootstrap": {
"draws": 19999,
"seed": 20261020,
"unit": "physician within each arm",
"order": [
"connected",
"comparison"
],
"rng": "PCG64 child stream per draw from SeedSequence",
"annual_records": "Retain all eligible records and physician multiplicities",
"weights": "Refit connected state-year masses and relative-entropy age/risk weights in every draw",
"CI": "95% percentile; linear quantile"
},
"historical_status": "Original 19999-draw bootstrap sampling frame and RNG order were not recovered; this is an explicit conditional analys
"interpretation": "The point estimate is approximately 25% higher after balancing. A lower nominal CI endpoint slightly above one is not a
}
}

#### *longitudinal_analysis.json*

{
"units": "2024 USD",
"event_study": {
"practice_mso": {
"coefficients": [
{
"bin": "le_minus4",
"label": "\u2264\u22124",
"event_time": -4,
"estimate": -20920.869130137868,
"se": 33663.05596585315,
"df": 13,
"ci": [
-93645.4801195198,
51803.74185924405
],
"p": 0.5450292613678717,
"treated_clusters": 14,
"clusters": 2108
},
{
"bin": "minus3",
"label": "\u22123",
"event_time": -3,
"estimate": 55717.18831483848,
"se": 64362.47023518442,
"df": 13,
"ci": [
-83329.47503377328,
194763.85166345024
],
"p": 0.402360405391409,
"treated_clusters": 14,
"clusters": 2108
},
{
"bin": "minus2",
"label": "\u22122",
"event_time": -2,
"estimate": 17749.460508969365,
"se": 22089.34627907172,
"df": 13,
"ci": [
-29971.670834090168,
65470.5918520289
],

"p": 0.4361148666501462,
"treated_clusters": 14,
"clusters": 2108
},
{
"bin": "plus1",
"label": "+1",
"event_time": 1,
"estimate": -6519.351875128132,
"se": 9139.019151579663,
"df": 13,
"ci": [
-26263.002401014004,
13224.298650757743
],
"p": 0.4882360802288282,
"treated_clusters": 14,
"clusters": 2108
},
{
"bin": "plus2",
"label": "+2",
"event_time": 2,
"estimate": -16256.592039824185,
"se": 14361.02557887502,
"df": 13,
"ci": [
-47281.70157508619,
14768.517495437822
],
"p": 0.2780859266875996,
"treated_clusters": 14,
"clusters": 2108
}
],
"reference_event_time": -1,
"excluded_event_time": 0,
"reference_estimated": false,
"units": "2024 USD",
"log_event_study": null,
"log_event_study_status": "not available in surviving JSON; not fabricated",
"counts": {
"treated_surgeons": 50,
"control_cell_units": 2801,
"distinct_controls": 2094,
"cells": 18,
"observations": 20023
}
},
"operator_owned": {
"coefficients": [
{
"bin": "le_minus4",
"label": "\u2264\u22124",
"event_time": -4,
"estimate": 43790.422740129296,
"se": 4945.450784321843,
"df": 2,
"ci": [
22511.865423125302,
65068.98005713329
],
"p": 0.012515276840481737,
"treated_clusters": 3,
"clusters": 1360
},
{
"bin": "minus3",
"label": "\u22123",
"event_time": -3,
"estimate": 5831.571132084848,
"se": 4140.203206182374,
"df": 2,
"ci": [
-11982.285494713216,
23645.427758882914
],
"p": 0.2943201801462251,
"treated_clusters": 3,
"clusters": 1360
},
{
"bin": "minus2",
"label": "\u22122",

```
        "event_time": -2,
        "estimate": -4822.878752838403,
        "se": 5989.392421382021,
        "df": 2,
        "ci": [
          -30593.154404238492,
          20947.39689856169
        ],
        "p": 0.5051981915521149,
        "treated_clusters": 3,
        "clusters": 1360
      },
      {
        "bin": "plus1",
        "label": "+1",
        "event_time": 1,
        "estimate": -14874.535248871422,
        "se": 4379.162223950583,
        "df": 2,
        "ci": [
          -33716.549545768125,
          3967.4790480252796
        ],
        "p": 0.07682059519696266,
        "treated_clusters": 3,
        "clusters": 1360
      },
      {
        "bin": "plus2",
        "label": "+2",
        "event_time": 2,
        "estimate": -20309.83711300476,
        "se": 5226.051881395452,
        "df": 2,
        "ci": [
          -42795.72350630321,
          2176.0492802936933
        ],
        "p": 0.06028622434796088,
        "treated_clusters": 3,
        "clusters": 1360
      }
    ],
    "reference_event_time": -1,
    "excluded_event_time": 0,
    "reference_estimated": false,
    "units": "2024 USD",
    "log_event_study": null,
    "log_event_study_status": "not available in surviving JSON; not fabricated",
    "counts": {
      "treated_surgeons": 20,
      "control_cell_units": 1387,
      "distinct_controls": 1357,
      "cells": 12,
      "observations": 8789
    }
  }
},
"practice_mso": {
  "counts": {
    "treated_surgeons": 50,
    "control_cell_units": 2801,
    "distinct_controls": 2094,
    "cells": 18,
    "observations": 20023
  },
  "levels": {
    "estimate": -15726.818352647058,
    "se": 28920.94188994502,
    "df": 13,
    "ci": [
      -78206.71472706604,
      46753.07802177192
    ],
    "p": 0.595791505391359,
    "treated_clusters": 14,
    "clusters": 2108,
    "wild_bootstrap": {
      "p": 0.8321,
      "ci": [
        -95893.46870100955,
        35190.739920870525
      ],
      "draws": 9999,
```

```
        "seed": 20261019,
        "weights": "Webb six-point",
        "null_imposed": true,
        "fixed_effects_refit_each_draw": [
          "cell_by_physician",
          "cell_by_year"
        ],
        "inversion": "two-sided restricted wild cluster bootstrap t; common draws across nulls",
        "monte_carlo_p": "(1+exceedances)/(draws+1)",
        "endpoint_tolerance_dollars": 0.01,
        "acceptance_grid_transitions": 2,
        "acceptance_grid_disconnected": false
      }
    },
    "log": {
      "estimate": 0.15654186352734728,
      "se": 0.07392204377343226,
      "df": 13,
      "ci": [
        -0.0031570028624462176,
        0.3162407299171408
      ],
      "p": 0.05404995410969808,
      "treated_clusters": 14,
      "clusters": 2108,
      "percent": 16.945971900321414,
      "percent_ci": [
        -0.3152024768905894,
        37.196048781061585
      ]
    }
  },
  "operator_owned": {
    "counts": {
      "treated_surgeons": 20,
      "control_cell_units": 1387,
      "distinct_controls": 1357,
      "cells": 12,
      "observations": 8789
    },
    "levels": {
      "estimate": -30847.726283344204
    },
    "log": {
      "percent": -13.85591448151361
    }
  },
  "sensitivities": [
    {
      "counts": {
        "treated_surgeons": 50,
        "control_cell_units": 2801,
        "distinct_controls": 2094,
        "cells": 18,
        "observations": 20023
      },
      "levels": {
        "estimate": -15726.818352647058,
        "se": 28920.94188994502,
        "df": 13,
        "ci": [
          -78206.71472706604,
          46753.07802177192
        ],
        "p": 0.595791505391359,
        "treated_clusters": 14,
        "clusters": 2108,
        "wild_bootstrap": {
          "p": 0.8321,
          "ci": [
            -95893.46870100955,
            35190.739920870525
          ],
          "draws": 9999,
          "seed": 20261019,
          "weights": "Webb six-point",
          "null_imposed": true,
          "fixed_effects_refit_each_draw": [
            "cell_by_physician",
            "cell_by_year"
          ],
          "inversion": "two-sided restricted wild cluster bootstrap t; common draws across nulls",
          "monte_carlo_p": "(1+exceedances)/(draws+1)",
          "endpoint_tolerance_dollars": 0.01,
```

```
      "acceptance_grid_transitions": 2,
      "acceptance_grid_disconnected": false
    }
  },
  "log": {
    "estimate": 0.15654186352734728,
    "se": 0.07392204377343226,
    "df": 13,
    "ci": [
      -0.0031570028624462176,
      0.3162407299171408
    ],
    "p": 0.05404995410969808,
    "treated_clusters": 14,
    "clusters": 2108,
    "percent": 16.945971900321414,
    "percent_ci": [
      -0.3152024768905894,
      37.196048781061585
    ]
  },
  "id": "full50"
},
{
  "counts": {
    "treated_surgeons": 46,
    "control_cell_units": 2554,
    "distinct_controls": 1848,
    "cells": 17,
    "observations": 18261
  },
  "levels": {
    "estimate": -19375.093726275372,
    "se": 31177.254704401013,
    "df": 13,
    "ci": [
      -86729.45758422044,
      47979.27013166969
    ],
    "p": 0.5450477324347237,
    "treated_clusters": 14,
    "clusters": 1862
  },
  "log": {
    "estimate": 0.1375613591679727,
    "se": 0.07488608392942696,
    "df": 13,
    "ci": [
      -0.024220189358403227,
      0.2993429076943487
    ],
    "p": 0.0891809365522137,
    "treated_clusters": 14,
    "clusters": 1862,
    "percent": 14.747211174607887,
    "percent_ci": [
      -2.392923430107486,
      34.89721170904809
    ]
  },
  "id": "sameZIP46"
},
{
  "counts": {
    "treated_surgeons": 49,
    "control_cell_units": 2801,
    "distinct_controls": 2094,
    "cells": 18,
    "observations": 20016
  },
  "levels": {
    "estimate": 8556.136814036001,
    "se": 10282.424910436746,
    "df": 13,
    "ci": [
      -13657.691674903768,
      30769.96530297577
    ],
    "p": 0.4203686905872869,
    "treated_clusters": 14,
    "clusters": 2108
  },
  "log": {
    "estimate": 0.18699534112239988,
```

```
        "se": 0.0674786052475824,
        "df": 13,
        "ci": [
          0.041216677363697246,
          0.33277400488110254
        ],
        "p": 0.015881701576356287,
        "treated_clusters": 14,
        "clusters": 2108,
        "percent": 20.562166819305645,
        "percent_ci": [
          4.207787577183699,
          39.483203761525544
        ]
      },
      "id": "99pct49"
    },
    {
      "counts": {
        "treated_surgeons": 47,
        "control_cell_units": 2518,
        "distinct_controls": 1813,
        "cells": 17,
        "observations": 18452
      },
      "levels": {
        "estimate": 21277.085377569783,
        "se": 9682.415496291396,
        "df": 12,
        "ci": [
          180.91427208130335,
          42373.25648305826
        ],
        "p": 0.0483533976825907,
        "treated_clusters": 13,
        "clusters": 1826
      },
      "log": {
        "estimate": 0.21152669374998506,
        "se": 0.07126122234733914,
        "df": 12,
        "ci": [
          0.05626182824183354,
          0.36679155925813656
        ],
        "p": 0.011737594221910645,
        "treated_clusters": 13,
        "clusters": 1826,
        "percent": 23.55629469968978,
        "percent_ci": [
          5.787462893316604,
          44.30970875359813
        ]
      },
      "id": "95pct47"
    }
  ],
  "leave_parent_out": [
    {
      "counts": {
        "treated_surgeons": 38,
        "control_cell_units": 2711,
        "distinct_controls": 2080,
        "cells": 17,
        "observations": 19465
      },
      "levels": {
        "estimate": -20793.924388269505,
        "se": 37190.077161485984,
        "df": 12,
        "ci": [
          -101824.14164402934,
          60236.29286749034
        ],
        "p": 0.5863596591728687,
        "treated_clusters": 13,
        "clusters": 2093
      },
      "log": {
        "estimate": 0.2058215673072823,
        "se": 0.0762801998958151,
        "df": 12,
        "ci": [
          0.03962128912469959,
```

```
        0.372021845489865
      ],
      "p": 0.019372676284792632,
      "treated_clusters": 13,
      "clusters": 2093,
      "percent": 22.853397381193524,
      "percent_ci": [
        4.041668246119408,
        45.06646714273915
      ]
    }
  },
  {
    "counts": {
      "treated_surgeons": 47,
      "control_cell_units": 2705,
      "distinct_controls": 1999,
      "cells": 17,
      "observations": 19158
    },
    "levels": {
      "estimate": -20565.25031582969,
      "se": 31095.732479391903,
      "df": 12,
      "ci": [
        -88317.0311898287,
        47186.530558169325
      ],
      "p": 0.5208825599960614,
      "treated_clusters": 13,
      "clusters": 2012
    },
    "log": {
      "estimate": 0.13941282190681928,
      "se": 0.076743390682062,
      "df": 12,
      "ci": [
        -0.027796662303421843,
        0.30662230611706043
      ],
      "p": 0.0943223875168429,
      "treated_clusters": 13,
      "clusters": 2012,
      "percent": 14.959858153718777,
      "percent_ci": [
        -2.741388988441465,
        35.88276502481139
      ]
    }
  },
  {
    "counts": {
      "treated_surgeons": 49,
      "control_cell_units": 2405,
      "distinct_controls": 2068,
      "cells": 17,
      "observations": 16842
    },
    "levels": {
      "estimate": -15685.800590378965,
      "se": 29108.26177040975,
      "df": 12,
      "ci": [
        -79107.25478505983,
        47735.65360430191
      ],
      "p": 0.5998258637426727,
      "treated_clusters": 13,
      "clusters": 2081
    },
    "log": {
      "estimate": 0.1585121426143856,
      "se": 0.07448824727879699,
      "df": 12,
      "ci": [
        -0.0037838062160823194,
        0.3208080914448535
      ],
      "p": 0.05474807581239065,
      "treated_clusters": 13,
      "clusters": 2081,
      "percent": 17.176615244335515,
      "percent_ci": [
        -0.3776656641719193,
```

```
      37.82410592622571
    ]
  }
},
{
  "counts": {
    "treated_surgeons": 42,
    "control_cell_units": 2704,
    "distinct_controls": 1997,
    "cells": 16,
    "observations": 19112
  },
  "levels": {
    "estimate": -19490.98313204557,
    "se": 35386.21307691931,
    "df": 12,
    "ci": [
      -96590.91817737561,
      57608.95191328447
    ],
    "p": 0.5918728228636981,
    "treated_clusters": 13,
    "clusters": 2010
  },
  "log": {
    "estimate": 0.1593872371363654,
    "se": 0.08956157496577338,
    "df": 12,
    "ci": [
      -0.035750671444264914,
      0.3545251457169957
    ],
    "p": 0.10045071984110765,
    "treated_clusters": 13,
    "clusters": 2010,
    "percent": 17.27920073789511,
    "percent_ci": [
      -3.511916415990583,
      42.5503587056315
    ]
  }
},
{
  "counts": {
    "treated_surgeons": 47,
    "control_cell_units": 2498,
    "distinct_controls": 2044,
    "cells": 17,
    "observations": 17736
  },
  "levels": {
    "estimate": -19042.340798782916,
    "se": 30925.497133952806,
    "df": 12,
    "ci": [
      -86423.21071807639,
      48338.52912051056
    ],
    "p": 0.5495612188849924,
    "treated_clusters": 13,
    "clusters": 2057
  },
  "log": {
    "estimate": 0.1494024007680839,
    "se": 0.07814921759461296,
    "df": 12,
    "ci": [
      -0.020870117155514734,
      0.3196749186916825
    ],
    "p": 0.08007702565809335,
    "treated_clusters": 13,
    "clusters": 2057,
    "percent": 16.114013889583028,
    "percent_ci": [
      -2.065384342619736,
      37.66801585986776
    ]
  }
},
{
  "counts": {
    "treated_surgeons": 48,
    "control_cell_units": 2658,
```

```
      "distinct_controls": 2031,
      "cells": 16,
      "observations": 19461
    },
    "levels": {
      "estimate": -15884.76349066006,
      "se": 29783.127858320444,
      "df": 12,
      "ci": [
        -80776.62457598808,
        49007.09759466797
      ],
      "p": 0.6035312177738617,
      "treated_clusters": 13,
      "clusters": 2044
    },
    "log": {
      "estimate": 0.16227007049675732,
      "se": 0.07607608640479618,
      "df": 12,
      "ci": [
        -0.003485482592885225,
        0.32802562358639986
      ],
      "p": 0.054264313493647964,
      "treated_clusters": 13,
      "clusters": 2044,
      "percent": 17.617784936684128,
      "percent_ci": [
        -0.3479415349569874,
        38.82245431252074
      ]
    }
  },
  {
    "counts": {
      "treated_surgeons": 43,
      "control_cell_units": 2571,
      "distinct_controls": 1867,
      "cells": 16,
      "observations": 18287
    },
    "levels": {
      "estimate": -20406.518656271714,
      "se": 33725.962904529224,
      "df": 12,
      "ci": [
        -93889.07932554101,
        53076.04201299758
      ],
      "p": 0.5564030117292935,
      "treated_clusters": 13,
      "clusters": 1880
    },
    "log": {
      "estimate": 0.1405762336223844,
      "se": 0.08330679051954826,
      "df": 12,
      "ci": [
        -0.040933670360007585,
        0.32208613760477633
      ],
      "p": 0.11731416605470958,
      "treated_clusters": 13,
      "clusters": 1880,
      "percent": 15.093681630320294,
      "percent_ci": [
        -4.010720282006846,
        38.00036410460183
      ]
    }
  },
  {
    "counts": {
      "treated_surgeons": 49,
      "control_cell_units": 2518,
      "distinct_controls": 1813,
      "cells": 17,
      "observations": 18469
    },
    "levels": {
      "estimate": -10685.045704181004,
      "se": 28663.245056774656,
      "df": 12,
```

```
      "ci": [
        -73136.89177377739,
        51766.80036541539
      ],
      "p": 0.7158142230669026,
      "treated_clusters": 13,
      "clusters": 1826
    },
    "log": {
      "estimate": 0.16401499433108307,
      "se": 0.07519840039736096,
      "df": 12,
      "ci": [
        0.0001717547748598114,
        0.32785823388730634
      ],
      "p": 0.04979591379163883,
      "treated_clusters": 13,
      "clusters": 1826,
      "percent": 17.82319817606991,
      "percent_ci": [
        0.017176952555564368,
        38.79921880841222
      ]
    }
  },
  {
    "counts": {
      "treated_surgeons": 48,
      "control_cell_units": 2513,
      "distinct_controls": 2060,
      "cells": 17,
      "observations": 18210
    },
    "levels": {
      "estimate": 11621.632169697437,
      "se": 10021.679667816776,
      "df": 12,
      "ci": [
        -10213.732065356962,
        33456.99640475184
      ],
      "p": 0.2687457502663206,
      "treated_clusters": 13,
      "clusters": 2073
    },
    "log": {
      "estimate": 0.19567526571823374,
      "se": 0.06901588471697606,
      "df": 12,
      "ci": [
        0.04530257064605189,
        0.34604796079041555
      ],
      "p": 0.015029426216901362,
      "treated_clusters": 13,
      "clusters": 2073,
      "percent": 21.61319214793334,
      "percent_ci": [
        4.634440512030745,
        41.347040534991386
      ]
    }
  },
  {
    "counts": {
      "treated_surgeons": 49,
      "control_cell_units": 2750,
      "distinct_controls": 2044,
      "cells": 17,
      "observations": 19735
    },
    "levels": {
      "estimate": -20047.217380592905,
      "se": 29539.756410276663,
      "df": 12,
      "ci": [
        -84408.81763254845,
        44314.38287136264
      ],
      "p": 0.5102349086231975,
      "treated_clusters": 13,
      "clusters": 2057
    },
```

```
    "log": {
      "estimate": 0.14834824103466118,
      "se": 0.07465069973167132,
      "df": 12,
      "ci": [
        -0.014301661284340211,
        0.3109981433536626
      ],
      "p": 0.07020897634888816,
      "treated_clusters": 13,
      "clusters": 2057,
      "percent": 15.991675664998706,
      "percent_ci": [
        -1.4199878326129625,
        36.478668725760485
      ]
    }
  },
  {
    "counts": {
      "treated_surgeons": 45,
      "control_cell_units": 2499,
      "distinct_controls": 1793,
      "cells": 16,
      "observations": 17851
    },
    "levels": {
      "estimate": -24513.058788684357,
      "se": 31845.824281326157,
      "df": 12,
      "ci": [
        -93899.14930416593,
        44873.03172679721
      ],
      "p": 0.45633875390728085,
      "treated_clusters": 13,
      "clusters": 1806
    },
    "log": {
      "estimate": 0.11448367925191284,
      "se": 0.06796339460660336,
      "df": 12,
      "ci": [
        -0.03359583686469107,
        0.26256319536851674
      ],
      "p": 0.11789542123149185,
      "treated_clusters": 13,
      "clusters": 1806,
      "percent": 12.129434055216032,
      "percent_ci": [
        -3.3037763838358623,
        30.025863589581636
      ]
    }
  },
  {
    "counts": {
      "treated_surgeons": 49,
      "control_cell_units": 2412,
      "distinct_controls": 2074,
      "cells": 17,
      "observations": 17117
    },
    "levels": {
      "estimate": -16758.442945527524,
      "se": 29532.011955557697,
      "df": 12,
      "ci": [
        -81103.16948018261,
        47586.28358912756
      ],
      "p": 0.5808583204474441,
      "treated_clusters": 13,
      "clusters": 2087
    },
    "log": {
      "estimate": 0.14241879146972153,
      "se": 0.07364354194229257,
      "df": 12,
      "ci": [
        -0.018036702536282173,
        0.30287428547572526
      ],
```

```
      "p": 0.07705820039867138,
      "treated_clusters": 13,
      "clusters": 2087,
      "percent": 15.30594388927976,
      "percent_ci": [
        -1.7875014781109582,
        35.374426842776195
      ]
    }
  },
  {
    "counts": {
      "treated_surgeons": 48,
      "control_cell_units": 2742,
      "distinct_controls": 2036,
      "cells": 17,
      "observations": 19467
    },
    "levels": {
      "estimate": -14805.109804101492,
      "se": 30214.873117442254,
      "df": 12,
      "ci": [
        -80637.66299915212,
        51027.443390949134
      ],
      "p": 0.6329739052419968,
      "treated_clusters": 13,
      "clusters": 2049
    },
    "log": {
      "estimate": 0.15280286706968949,
      "se": 0.07705015506584285,
      "df": 12,
      "ci": [
        -0.015074999315618315,
        0.32068073345499726
      ],
      "p": 0.07071120113100966,
      "treated_clusters": 13,
      "clusters": 2049,
      "percent": 16.509527765646258,
      "percent_ci": [
        -1.4961940347707494,
        37.806554042851225
      ]
    }
  },
  {
    "counts": {
      "treated_surgeons": 48,
      "control_cell_units": 2727,
      "distinct_controls": 2022,
      "cells": 17,
      "observations": 19389
    },
    "levels": {
      "estimate": -15903.359627099886,
      "se": 30040.159253545055,
      "df": 12,
      "ci": [
        -81355.24401397056,
        49548.52475977079
      ],
      "p": 0.6061811316221603,
      "treated_clusters": 13,
      "clusters": 2035
    },
    "log": {
      "estimate": 0.1629196099270976,
      "se": 0.07666576796480237,
      "df": 12,
      "ci": [
        -0.004120748910904637,
        0.3299599687650998
      ],
      "p": 0.05503666928386557,
      "treated_clusters": 13,
      "clusters": 2035,
      "percent": 17.694207142640963,
      "percent_ci": [
        -0.41122702752189905,
        39.09124474063893
      ]
```

```
      }
    }
  ]
}
```


##### *aggregate_timing_audit.json*


```
{
  "origin": "Current manuscript source-documentation and timing-sensitivity section",
  "candidate_documentary_records": 10474,
  "accepted_before_deduplication": 3046,
  "rejected": 2130,
  "unresolved": 5298,
  "duplicates_removed": 34,
  "eligible_dated_observations": 3012,
  "distinct_connected_surgeons": 104,
  "practice_mso_surgeons": 69,
  "hospital_operator_surgeons": 35,
  "index_year_cms_snapshot_affiliations_with_matching_organization": 81,
  "remaining_surgeons": 23,
  "remaining_with_retained_in_window_document": 17,
  "remaining_with_in_window_cms_affiliation": 5,
  "total_with_investment_window_corroboration": 103,
  "calendar_year_supported_index_assignments": 102,
  "uncertain_index_assignments": 2,
  "timing_exceptions_longitudinally_eligible": 0,
  "scope": "Aggregate audit only. No source inventories, original documents, or linkage key."
}
```


##### *VALIDATION.json*


```
{
  "table_count": 12,
  "source_urls": 27,
  "organization_rows": 23,
  "numerical_checks": [
    {
      "result": "state_year_ratio",
      "rounded": [
        1.31,
        1.07,
        1.6
      ],
      "matched_current_manuscript": true
    },
    {
      "result": "balanced_ratio",
      "rounded": [
        1.25,
        1.01,
        1.59
      ],
      "matched_current_manuscript": true
    },
    {
      "result": "balanced_dollar_difference",
      "rounded": [
        19708.0,
        -10419.0,
        52919.0
      ],
      "matched_current_manuscript": true
    }
  ],
  "wild_bootstrap_ci_matched": true,
  "scope": "Selected saved-output checks plus manuscript extraction; not a full independent reanalysis."
}
```

#### 4. Bibliography and Timing Audit Notes

##### *investment_bibliography/README.txt*

The 23 organization rows reproduce Table S1. The 27 URLs were recovered from its
supplemental reference hyperlinks, not guessed or newly substituted.
Dates are analytical eligibility windows, not universally verified holding periods.
Multiple starts can reflect different affiliates within a parent label.
Sources support investment relationships, not individual physician employment.
URLs were extracted on 2026-11-09; live availability was not reverified.
Counts of one identify small organizational groups, but no individual records are included.

##### *timing_audit/README.txt*

Investment-window corroboration (103/104) and index-calendar-year support (102/104)
are different questions and must not be conflated.
Table S9 is in aggregate_results/Table_S9.csv. P074 and P080 are the manuscript's
public study IDs; the package contains no mapping to names or NPIs.
P074's retained roster is from the following year and 33 days after the recorded
window ended; P080's roster is in-window but from the following calendar year.
Both sensitivity exclusions retain the original 4,016-person comparison pool.
Neither exception enters a longitudinal model. Source counts were exported from
the manuscript, not re-adjudicated during packaging.

Bibliography CSV tables (sources.csv, organization_windows.csv) are in Supplementary_Data_1.xlsx sheets named sources and organization_windows.

#### 5. Analytical Code (full source)

Every file from the code/ directory is reproduced below in full, unchanged.

##### *code/CODE_PROVENANCE.json*


```
{
  "alternative_tests.py": {
    "original_sha256": "cafd830cb040b32feda5e79b76d0dab7952977f447df4a8d1b4b633e50e56f50",
    "policy": "Function/class bodies extracted without modification; imports/constants curated; historical mains omitted."
  },
  "longitudinal_did.py": {
    "original_sha256": "c4c62811e3adaac868c3102e1899bff09eefe5907a821a1aa7496a3855110a6c",
    "policy": "Function/class bodies extracted without modification; imports/constants curated; historical mains omitted."
  },
  "positive_balance.py": {
    "original_sha256": "ff2872c67d06cf73629aec35538c031d92453088d0e1f10309adf9051f85a1c4",
    "policy": "Function/class bodies extracted without modification; imports/constants curated; historical mains omitted."
  },
  "primary_analysis.py": {
    "original_sha256": "a49ba79262a0cbc33187914416ca1ed18021298fb790510e34ca4e9f4cfe793c",
    "policy": "Function/class bodies extracted without modification; imports/constants curated; historical mains omitted."
  },
  "publication_balance.py": {
    "original_sha256": "e111cc98d61d0b93de6ece6b3cb99e418f0833c0781e330192fe6ed7ff7f9929",
    "policy": "Function/class bodies extracted without modification; imports/constants curated; historical mains omitted."
  },
  "publication_longitudinal.py": {
    "original_sha256": "9f66383b17315e314318797157ab5193cec4fbd10cba4d8d0ac9c00ae183bd26",
    "policy": "Function/class bodies extracted without modification; imports/constants curated; historical mains omitted."
  }
}
```


##### *code/INPUT_SCHEMA.txt*

```
INPUTS REQUIRED FOR FULL ANALYSIS (NOT INCLUDED IN THE PUBLIC ZIP)

Identifiers: the historical code uses a column called npi as a within-dataset
physician key. Consistent pseudonymous keys may be substituted throughout.
No linkage key belongs in this public package. CSVs must be UTF-8.

analysis_rows.csv
  npi, year, state, group, payment_real, bene_age, bene_risk
  group is connected or comparison. Each connected physician has one index
  record; comparisons can have repeated eligible years.
  payment_real is positive annual Medicare professional payment in 2024 USD.
  bene_age and bene_risk are provider-level means, not patient-level records.
  Required cohort: 104 connected (69 practice/MSO, 35 hospital operator);
  4,016 comparisons, 8,118 comparison annual records.

professional_index.csv
  npi, connection_type
  One row per connected physician. Allowed connection types:
  practice_or_linked_mso; operator_professional_group.

Additional files needed with --longitudinal:

national_neurosurgery_panel.csv
  npi, year, Rndrng_Prvdr_Ent_Cd, Rndrng_Prvdr_Type,
  Rndrng_Prvdr_State_Abrvtn, Tot_Mdcr_Pymt_Amt
  Payments here are NOMINAL annual amounts. Code uses CPI-U below.
  Entity I; specialty neurosurgery; one physician-year row.
  Missing/suppressed values must not be replaced with zero.

cpi_annual_2013_2024.csv
  year, cpi_u_annual
  Annual all-items CPI-U; positive values; includes 2024.

comparator_exclusions.csv
  npi
  Complete exclusion inventory, including connected physicians and known/
  unresolved connections. This is not the set of people omitted in a
  sensitivity specification.

longitudinal_eligibility.csv
  npi, connection_type, operator_parent, state, investment_start,
  cms_pre_and_post_same_state, plus_same_zip_before_investment
  Investment date YYYY-MM-DD; flags True or False.
  Eligible counts: 50 practice/MSO and 20 hospital operator.
  Parent labels must be consistent because they define treated clusters.

Other original metadata fields are not necessary for the generic driver.
Private input reconstruction, exposure verification, RUCA retrieval, and
```

source-timing adjudication are separate from these analytical routines.

##### code/requirements.txt

```
numpy>=2.0,<3
scipy>=1.14,<2
```

##### code/run_analysis.py

```
"""Run public analytical routines on separately obtained physician-level inputs."""
import argparse
import csv
import json
from pathlib import Path

import numpy as np
import alternative_tests as tests
import publication_balance as pb
import longitudinal_did as did
import publication_longitudinal as pl


def rows(path):
    with path.open(newline="", encoding="utf-8") as stream:
        return list(csv.DictReader(stream))


def save(path, obj):
    path.write_text(json.dumps(obj, indent=2, allow_nan=False) + "\n")


def main():
    parser = argparse.ArgumentParser(description=__doc__)
    parser.add_argument("--input-dir", required=True, type=Path)
    parser.add_argument("--output-dir", required=True, type=Path)
    parser.add_argument("--draws", type=int, default=19999)
    parser.add_argument("--wild-draws", type=int, default=9999)
    parser.add_argument("--longitudinal", action="store_true")
    args = parser.parse_args()
    if min(args.draws, args.wild_draws) < 2:
        parser.error("At least two draws required")
    args.output_dir.mkdir(parents=True, exist_ok=True)
    records = rows(args.input_dir / "analysis_rows.csv")
    index = rows(args.input_dir / "professional_index.csv")
    kinds = {r["npi"]: r["connection_type"] for r in index}
    means = tests.physician_means(records)
    controls = np.array([r["mean_annual_payment_2024_usd"] for r in means if r["group"] == "comparison"])
    result = {}
    for name, kind in (("practice_mso", "practice_or_linked_mso"),
                       ("operator_owned", "operator_professional_group"), ("pooled", None)):
        exposed = np.array([r["mean_annual_payment_2024_usd"] for r in means
                            if r["group"] == "connected" and (kind is None or kinds[r["npi"]] == kind)])
        connected_records = [r for r in records if r["group"] == "connected"
                             and (kind is None or kinds[r["npi"]] == kind)]
        annual_controls = [(r["state"] + ":" + r["year"], float(r["payment_real"]))
                           for r in records if r["group"] == "comparison"]
        result[name] = {
            "dollar": tests.welch(exposed, controls), "log_ratio": tests.log_ratio(exposed, controls),
            "median_point_and_rank_p": tests.median_comparison(exposed, controls),
            "state_year_dollar_point": tests.standardized(
                [(r["state"] + ":" + r["year"], float(r["payment_real"])) for r in connected_records],
                annual_controls)}
    save(args.output_dir / "cross_sectional.json", result)
    data = pb.prepare(records, index)
    point, weights, base, diagnostics = pb.estimate(data)
    assert (len(data["ty"]), data["ncontrol"], len(data["cy"]), len(data["strata"])) == (69, 3294, 5943, 36)
    boot = []
    for i in range(args.draws):
        tc, cc = pb.draw_counts(data, i)
        values, _, _, _ = pb.estimate(data, tc, cc)
        boot.append(values)
        if (i + 1) % 1000 == 0:
            print(f"Weighting bootstrap {i+1}/{args.draws}", flush=True)
    summaries = pb.summarize_bootstrap(np.array(boot))
    for j, key in enumerate(pb.MEASURES):
        summaries[key]["estimate"] = float(point[j])
    save(args.output_dir / "weighted_analysis.json", {
        "seed": pb.SEED, "draws": args.draws, "estimates": summaries,
        "balance_diagnostics": diagnostics,
        "concentration": pb.concentration(weights, base, data["person"])})
    if not args.longitudinal:
        return
    did.DATA = args.input_dir
    did.PANEL = args.input_dir / "national_neurosurgery_panel.csv"
```

```
    panel = did.load_panel()
    exclusions = {r["npi"] for r in rows(args.input_dir / "comparator_exclusions.csv")}
    eligible = [r for r in rows(args.input_dir / "longitudinal_eligibility.csv")
                if r["cms_pre_and_post_same_state"] == "True"]
    out, stacks, cohort = {}, {}, {}
    for arm, kind in (("practice_mso", "practice_or_linked_mso"), ("operator_owned", "operator_professional_group")):
        cohort[arm] = [r for r in eligible if r["connection_type"] == kind]
        stack = did.build_stack(pl.treated_rows(cohort[arm]), panel, exclusions)
        stacks[arm] = stack
        pair = pl.model_pair(stack)
        if arm == "operator_owned":
            pair["levels"] = {"estimate": pair["levels"]["estimate"]}
            pair["log"] = {"percent": pair["log"]["percent"]}
        et, treated = stack["event_time"], stack["treated"] == 1
        bins = {"le_minus4": et <= -4, "minus3": et == -3, "minus2": et == -2,
                "plus1": et == 1, "plus2": et == 2}
        columns = [(treated & mask).astype(float) for mask in bins.values()]
        keep = [k for k, col in zip(bins, columns) if col.sum() > 0]
        model = did.fit(stack, [col for col in columns if col.sum() > 0], stack["y"])
        pair["event_study"] = {k: did.summarize(model, i) for i, k in enumerate(keep)}
        pair["event_study"]["reference"] = "minus1"
        pair.pop("parents", None)
        out[arm] = pair
    moments = pl.bootstrap_moments(stacks["practice_mso"], draws=args.wild_draws)
    out["practice_mso"]["levels"]["wild_bootstrap"] = pl.invert_bootstrap(moments)
    practice = cohort["practice_mso"]
    pre = pl.preinvestment_means(practice, panel)
    sens = {}
    for label, quantile in (("above_99_removed", .99), ("above_95_removed", .95)):
        cutoff = np.quantile(list(pre.values()), quantile)
        selected = [r for r in practice if pre[r["npi"]] <= cutoff]
        value = pl.model_pair(did.build_stack(pl.treated_rows(selected), panel, exclusions))
        value.pop("parents", None)
        sens[label] = value
    selected = [r for r in practice if r["plus_same_zip_before_investment"] == "True"]
    value = pl.model_pair(did.build_stack(pl.treated_rows(selected), panel, exclusions))
    value.pop("parents", None)
    sens["same_zip"] = value
    out["sensitivities"] = sens
    out["leave_parent_out"] = {}
    for parent in sorted({r["operator_parent"] for r in practice}):
        selected = [r for r in practice if r["operator_parent"] != parent]
        value = pl.model_pair(did.build_stack(pl.treated_rows(selected), panel, exclusions))
        value.pop("parents", None)
        out["leave_parent_out"][parent] = value
    save(args.output_dir / "longitudinal_analysis.json", out)


if __name__ == "__main__":
    main()
```

##### *code/primary_analysis.py*

```
"""Public algorithm extract. See README and CODE_PROVENANCE.json."""
import math
from collections import defaultdict

def physician_means(rows):
    grouped, seen, arms = defaultdict(list), set(), {}
    for row in rows:
        npi, year, group = row["npi"], row["year"], row["group"]
        if group not in {"connected", "comparison"}:
            raise ValueError("Unknown group")
        if (npi, year) in seen:
            raise ValueError("Duplicate NPI-year")
        if npi in arms and arms[npi] != group:
            raise ValueError("NPI occurs in both groups")
        seen.add((npi, year))
        arms[npi] = group
        value = float(row["payment_real"])
        if not math.isfinite(value):
            raise ValueError("Missing/nonfinite payment; do not silently exclude")
        grouped[group, npi].append(value)
    if any(len(values) != 1 for (group, _), values in grouped.items()
           if group == "connected"):
        raise ValueError("Connected surgeon has multiple index records")
    return [
        {"group": group, "npi": npi, "eligible_years": len(values),
         "mean_annual_payment_2024_usd": math.fsum(values)/len(values)}
        for (group, npi), values in sorted(grouped.items())
    ]
```

##### *code/alternative_tests.py*

```
"""Public algorithm extract. See README and CODE_PROVENANCE.json."""
import math
from collections import Counter, defaultdict
import numpy as np
from scipy import stats
from positive_balance import balance
from primary_analysis import physician_means

def welch(a, b):
    a, b = np.asarray(a, float), np.asarray(b, float)
    va, vb = a.var(ddof=1) / len(a), b.var(ddof=1) / len(b)
    se = math.sqrt(va + vb)
    df = (va + vb) ** 2 / (va ** 2 / (len(a) - 1) + vb ** 2 / (len(b) - 1))
    diff = float(a.mean() - b.mean())
    crit = stats.t.ppf(0.975, df)
    p = float(2 * stats.t.sf(abs(diff / se), df))
    check = stats.ttest_ind(a, b, equal_var=False)
    assert math.isclose(p, check.pvalue, rel_tol=1e-9, abs_tol=1e-15)
    return {"n_connected": len(a), "n_comparison": len(b), "connected_mean": float(a.mean()),
            "comparison_mean": float(b.mean()), "difference": diff, "se": se, "df": df,
            "ci": [diff - crit * se, diff + crit * se], "p": p}

def log_ratio(a, b):
    out = welch(np.log(a), np.log(b))
    return {"n_connected": out["n_connected"], "n_comparison": out["n_comparison"],
            "geometric_mean_connected": math.exp(out["connected_mean"]),
            "geometric_mean_comparison": math.exp(out["comparison_mean"]),
            "ratio": math.exp(out["difference"]), "ci": [math.exp(x) for x in out["ci"]],
            "df": out["df"], "p": out["p"]}

def median_comparison(a, b):
    return {"median_connected": float(np.median(a)), "median_comparison": float(np.median(b)),
            "difference": float(np.median(a) - np.median(b)),
            "mann_whitney_p": float(stats.mannwhitneyu(a, b, alternative="two-sided").pvalue)}

def standardized(connected, comparison_rows):
    """connected: list of (stratum, payment); comparison_rows: list of (stratum, payment)."""
    cells = defaultdict(list)
    for s, y in comparison_rows:
        cells[s].append(y)
    counts = Counter(s for s, _ in connected)
    exposed = defaultdict(list)
    for s, y in connected:
        exposed[s].append(y)
    n = len(connected)
    return math.fsum(counts[s] / n * (np.mean(exposed[s]) - np.mean(cells[s])) for s in counts)

def balanced(connected, comparison_rows, tol=1e-8):
    """connected/comparison: lists of (stratum, payment, age, risk)."""
    masses = {s: c / len(connected) for s, c in Counter(r[0] for r in connected).items()}
    rows = [r for r in comparison_rows if r[0] in masses]
    z = np.array([[r[2], r[3]] for r in rows])
    target = np.array([[r[2], r[3]] for r in connected]).mean(axis=0)
    weights, _, _ = balance(z, np.array([r[0] for r in rows]), masses, target, tol=tol)
    return float(np.mean([r[1] for r in connected]) - weights @ np.array([r[1] for r in rows]))
```

##### *code/positive_balance.py*

```
"""Public algorithm extract. See README and CODE_PROVENANCE.json."""
import numpy as np
from scipy.optimize import minimize

def balance(control_covariates, control_strata, target_masses, target_mean, tol=1e-8):
    z = np.asarray(control_covariates, dtype=float)
    target = np.asarray(target_mean, dtype=float)
    codes = np.asarray(control_strata)
    if (z.ndim != 2 or not len(z) or not z.shape[1]
            or target.shape != (z.shape[1],) or codes.shape != (len(z),)):
        raise ValueError("Invalid covariate, target, or stratum dimensions")
    if not np.isfinite(z).all() or not np.isfinite(target).all():
        raise ValueError("Covariates and targets must be finite")
    if set(codes) != set(target_masses):
        raise ValueError("Control strata and target strata must match")
    keys = sorted(target_masses)
    masses = np.array([target_masses[key] for key in keys], dtype=float)
    if not np.isfinite(masses).all() or np.any(masses <= 0):
        raise ValueError("Target masses must be positive and finite")
    if not np.isclose(masses.sum(), 1, rtol=0, atol=1e-12):
        raise ValueError("Target mass must sum to one")
    base = np.zeros(len(z))
    for key in keys:
        cell = codes == key
        if not cell.any() or target_masses[key] <= 0:
            raise ValueError("Unsupported target")
```

```
        base[cell] = target_masses[key] / cell.sum()
    if not np.isclose(base.sum(), 1, rtol=0, atol=1e-12):
        raise ValueError("Target mass must sum to one")
    center = base @ z
    scale = np.sqrt(base @ ((z - center) ** 2))
    if np.any(scale <= 0):
        raise ValueError("Degenerate covariates")
    zs, ts = (z - center) / scale, (target - center) / scale
    cells = [(codes == key, target_masses[key]) for key in keys]

    def evaluate(beta):
        value, gradient = -beta @ ts, -ts.copy()
        hessian = np.zeros((len(beta), len(beta)))
        weights = np.zeros(len(z))
        for cell, mass in cells:
            x = zs[cell]
            scores = x @ beta
            lognorm = logsumexp(scores)
            probabilities = np.exp(scores - lognorm)
            mean = probabilities @ x
            value += mass * (lognorm - np.log(len(x)))
            gradient += mass * mean
            deviation = x - mean
            hessian += mass * (deviation.T @ (probabilities[:, None] * deviation))
            weights[cell] = mass * probabilities
        return value, gradient, hessian, weights

    result = minimize(lambda b: evaluate(b)[0], np.zeros(z.shape[1]),
                      jac=lambda b: evaluate(b)[1],
                      hess=lambda b: evaluate(b)[2], method="trust-exact",
                      options={"gtol": 1e-10, "maxiter": 200})
    _, gradient, _, weights = evaluate(result.x)
    if (not np.isfinite(gradient).all() or not np.isfinite(weights).all()
            or np.max(abs(gradient)) > tol or not np.all(weights > 0)):
        raise ValueError(f"Balance failed: {result.message}; gradient={gradient}")
    direct_error = (weights @ z - target) / scale
    if np.max(abs(direct_error)) > tol:
        raise ValueError("Direct covariate balance failed")
    for cell, mass in cells:
        if abs(weights[cell].sum() - mass) > 1e-10:
            raise ValueError("Stratum mass changed")
    return weights, base, {
        "solver": "scipy trust-exact, convex entropy dual",
        "max_standardized_balance_error": float(max(abs(gradient))),
        "max_direct_standardized_balance_error": float(max(abs(direct_error))),
        "positivity_interpretation": "Numerically positive weights at the balance tolerance; not a certificate of strict-interior feasibility o
        "covariate_means": (weights @ z).tolist(),
        "target_means": target.tolist(),
        "minimum_weight": float(min(weights)),
        "maximum_weight": float(max(weights)),
        "max_ratio_to_state_year_weight": float(max(weights / base)),
        "comparison_weight_effective_n": float(1 / (weights @ weights)),
    }
```

#### *code/publication_balance.py*

```
"""Public algorithm extract. See README and CODE_PROVENANCE.json."""
import numpy as np
from scipy.optimize import minimize
TOL = 1e-8
SEED = 20261020
MEASURES = ("state_year_ratio","balanced_ratio","balanced_dollar_difference")

def prepare(rows, index):
    """Select comparison annual records in the 36 treated state-year strata."""
    kinds = {r["npi"]: r["connection_type"] for r in index}
    treated = sorted((r for r in rows if r["group"] == "connected" and
                      kinds[r["npi"]] == "practice_or_linked_mso"),
                     key=lambda r: (r["npi"], r["year"]))
    key = lambda r: r["state"] + ":" + r["year"]
    strata = sorted({key(r) for r in treated})
    code = {s: i for i, s in enumerate(strata)}
    controls = sorted((r for r in rows if r["group"] == "comparison" and key(r) in code),
                      key=lambda r: (r["npi"], r["year"]))
    if not treated or not controls:
        raise ValueError("Empty cohort")
    seen, arms = set(), {}
    for r in treated + controls:
        pair = r["npi"], r["year"]
        if pair in seen or (r["npi"] in arms and arms[r["npi"]] != r["group"]):
            raise ValueError("Duplicate physician-year or overlapping groups")
        seen.add(pair)
        arms[r["npi"]] = r["group"]
    if len({r["npi"] for r in treated}) != len(treated):
```

```python
        raise ValueError("Treated physicians require one index-year record")
    npis = sorted({r["npi"] for r in controls})
    people = {n: i for i, n in enumerate(npis)}
    cov = lambda records: np.array([[float(r[k]) for k in ("bene_age", "bene_risk")]
                                    for r in records])
    outcome = lambda records: np.array([float(r["payment_real"]) for r in records])
    data = {"tz": cov(treated), "cz": cov(controls), "ty": outcome(treated),
            "cy": outcome(controls), "ts": np.array([code[key(r)] for r in treated]),
            "cs": np.array([code[key(r)] for r in controls]),
            "person": np.array([people[r["npi"]] for r in controls]),
            "ncontrol": len(npis), "strata": strata}
    if any(not np.isfinite(data[k]).all() for k in ("tz", "cz", "ty", "cy")):
        raise ValueError("Nonfinite outcome/covariate; no silent exclusions")
    if np.any(data["ty"] <= 0) or np.any(data["cy"] <= 0):
        raise ValueError("Log payment requires strictly positive outcomes")
    return data

def entropy_weights(z, codes, masses, target, multiplicity=None, tol=TOL):
    """Frequency-aware convex dual using vectorized segmented moments.

    The returned weight is the TOTAL weight of all copies of a record. Zero
    copies and zero-target strata have zero weight. This is exactly equivalent
    to physically expanding every selected physician's annual records.
    """
    z, codes = np.asarray(z, float), np.asarray(codes, int)
    masses, target = np.asarray(masses, float), np.asarray(target, float)
    mult = np.ones(len(z)) if multiplicity is None else np.asarray(multiplicity, float)
    if (z.ndim != 2 or z.shape[1] != 2 or codes.shape != (len(z),) or
            mult.shape != (len(z),) or target.shape != (2,) or masses.ndim != 1):
        raise ValueError("Invalid dimensions")
    if (not all(np.isfinite(a).all() for a in (z, mult, masses, target)) or
            np.any(mult < 0) or np.any(masses < 0) or
            not np.isclose(masses.sum(), 1, atol=1e-12, rtol=0) or
            np.any(codes < 0) or np.any(codes >= len(masses))):
        raise ValueError("Invalid finite masses, multiplicities, or codes")
    counts = np.bincount(codes, weights=mult, minlength=len(masses))
    if np.any((masses > 0) & (counts == 0)):
        raise ValueError("Unsupported target stratum")
    active = (mult > 0) & (masses[codes] > 0)
    s, x, frequency = codes[active], z[active], mult[active]
    base = masses[s] * frequency / counts[s]
    center = base @ x
    scale = np.sqrt(base @ ((x - center) ** 2))
    if np.any(scale <= 0):
        raise ValueError("Degenerate covariates")
    x = (x - center) / scale
    target_s = (target - center) / scale
    logfrequency = np.log(frequency)
    cells = len(masses)

    def evaluate(beta):
        scores = x @ beta + logfrequency
        maxima = np.full(cells, -np.inf)
        np.maximum.at(maxima, s, scores)
        e = np.exp(scores - maxima[s])
        total = np.bincount(s, weights=e, minlength=cells)
        w = masses[s] * e / total[s]
        active_cells = masses > 0
        value = (masses[active_cells] @ (maxima[active_cells] +
                 np.log(total[active_cells]) - np.log(counts[active_cells])) - beta @ target_s)
        mean = w @ x
        g = mean - target_s
        # Subtract each stratum's mean product, not the global mean product.
        mu = np.column_stack([np.bincount(s, weights=w*x[:, j], minlength=cells)
                              for j in range(2)])
        h = x.T @ (w[:, None] * x)
        h -= (mu[active_cells].T / masses[active_cells]) @ mu[active_cells]
        return float(value), g, h, w

    beta = np.zeros(2)
    converged = False
    iterations = 0
    for iterations in range(1, 61):
        value, g, h, w = evaluate(beta)
        if np.max(np.abs(g)) <= tol * .1:
            converged = True
            break
        try:
            step = np.linalg.solve(h, g)
        except np.linalg.LinAlgError:
            break
        slope = g @ step
        if not np.isfinite(step).all() or slope <= 0:
            break
```

```
        length = 1.
        for _ in range(35):
            candidate = beta - length * step
            trial = evaluate(candidate)
            # Also accept a strictly improving gradient near machine precision.
            if (trial[0] <= value - 1e-4 * length * slope or
                    (abs(trial[0] - value) < 1e-14 and
                     np.linalg.norm(trial[1]) < np.linalg.norm(g))):
                beta = candidate
                break
            length *= .5
        else:
            break
    fallback = False
    if not converged:
        fallback = True
        result = minimize(lambda b: evaluate(b)[0], beta,
                          jac=lambda b: evaluate(b)[1], hess=lambda b: evaluate(b)[2],
                          method="trust-exact", options={"gtol": tol*.1, "maxiter": 200})
        beta = result.x
    _, g, _, w = evaluate(beta)
    direct = (w @ z[active] - target) / scale
    mass_error = np.max(np.abs(np.bincount(s, weights=w, minlength=cells) - masses))
    error = float(max(np.max(np.abs(g)), np.max(np.abs(direct))))
    if (error > tol or mass_error > 1e-10 or not np.isfinite(w).all() or np.any(w <= 0)):
        raise ValueError(f"Entropy convergence failed: error={error}, mass_error={mass_error}")
    weights, original = np.zeros(len(z)), np.zeros(len(z))
    weights[active], original[active] = w, base
    return weights, original, {"max_standardized_balance_error": error,
                               "max_stratum_mass_error": float(mass_error),
                               "iterations": iterations, "fallback": fallback,
                               "target_means": target.tolist(),
                               "weighted_means": (weights @ z).tolist()}

def estimate(data, treated_counts=None, control_counts=None):
    nt = len(data["ty"])
    tc = np.ones(nt) if treated_counts is None else np.asarray(treated_counts)
    cc = np.ones(data["ncontrol"]) if control_counts is None else np.asarray(control_counts)
    if tc.shape != (nt,) or cc.shape != (data["ncontrol"],) or tc.sum() <= 0:
        raise ValueError("Invalid physician multiplicities")
    masses = np.bincount(data["ts"], weights=tc, minlength=len(data["strata"])) / tc.sum()
    target = tc @ data["tz"] / tc.sum()
    record_counts = cc[data["person"]]
    w, base, diagnostics = entropy_weights(data["cz"], data["cs"], masses,
                                           target, record_counts)
    log_t = float(tc @ np.log(data["ty"]) / tc.sum())
    log_c = np.log(data["cy"])
    values = np.array([np.exp(log_t - base @ log_c), np.exp(log_t - w @ log_c),
                       tc @ data["ty"] / tc.sum() - w @ data["cy"]])
    return values, w, base, diagnostics

def draw_counts(data, draw, seed=SEED):
    # A separate deterministic SeedSequence stream for every draw: scheduling,
    # worker count, failures, and requested draw count cannot shift later draws.
    rng = np.random.default_rng(np.random.SeedSequence(seed, spawn_key=(int(draw),)))
    nt, nc = len(data["ty"]), data["ncontrol"]
    tc = np.bincount(rng.integers(nt, size=nt), minlength=nt)
    cc = np.bincount(rng.integers(nc, size=nc), minlength=nc)
    return tc, cc

def concentration(weights, base, person):
    physician = np.bincount(person, weights=weights)
    ordered = np.sort(weights)[::-1]
    return {"minimum_record_weight": float(weights.min()),
            "maximum_record_weight": float(weights.max()),
            "maximum_ratio_to_state_year_weight": float(np.max(weights/base)),
            "record_effective_n": float(1/(weights@weights)),
            "physician_effective_n": float(1/(physician@physician)),
            "maximum_physician_weight": float(physician.max()),
            "top_1_percent_record_mass": float(ordered[:int(np.ceil(.01*len(weights)))].sum()),
            "top_10_record_mass": float(ordered[:10].sum())}

def summarize_bootstrap(values):
    summaries = {}
    for j, name in enumerate(MEASURES):
        v = values[:, j]
        summaries[name] = {"ci": np.quantile(v, [.025, .975], method="linear").tolist(),
                           "bootstrap_mean": float(v.mean()),
                           "bootstrap_sd": float(v.std(ddof=1)),
                           "prefix_convergence": []}
        for n in (999, 4999, 9999, 14999, 19999):
            if n <= len(v):
                summaries[name]["prefix_convergence"].append(
                    {"draws": n, "ci": np.quantile(v[:n], [.025, .975]).tolist()})
```

```
        # Distribution-free order-statistic uncertainty of percentile endpoints.
        # Normal binomial rank approximation: 95% MC range for each quantile.
        mc = []
        for p in (.025, .975):
            width = 1.96*np.sqrt(p*(1-p)/len(v))
            mc.append(np.quantile(v, [max(0, p-width), min(1, p+width)]).tolist())
        summaries[name]["endpoint_monte_carlo_95_ranges"] = mc
    return summaries
```

#### *code/longitudinal_did.py*

```
"""Public algorithm extract. See README and CODE_PROVENANCE.json."""
import csv, math
from collections import defaultdict
from pathlib import Path
import numpy as np
from scipy import stats
DATA = Path(__file__).resolve().parent.parent / 'private_inputs'
PANEL = DATA / 'national_neurosurgery_panel.csv'
EXCLUSIONS = DATA / 'comparator_exclusions.csv'
POST_YEARS = 2
SEED = 20261019
WEBB = np.array([-math.sqrt(1.5),-1.,-math.sqrt(.5),math.sqrt(.5),1.,math.sqrt(1.5)])

def payment(value):
    value = (value or "").replace(",", "").strip()
    return float(value) if value else None

def load_panel():
    cpi = {int(r["year"]): float(r["cpi_u_annual"]) for r in csv.DictReader((DATA / "cpi_annual_2013_2024.csv").open())}
    records = defaultdict(dict)
    with PANEL.open(newline="") as stream:
        for row in csv.DictReader(stream):
            if row["Rndrng_Prvdr_Ent_Cd"] != "I" or row["Rndrng_Prvdr_Type"].lower() != "neurosurgery":
                continue
            year, value = int(row["year"]), payment(row["Tot_Mdcr_Pymt_Amt"])
            assert year not in records[row["npi"]], "duplicate physician-year"
            records[row["npi"]][year] = {
                "state": row["Rndrng_Prvdr_State_Abrvtn"],
                "y": None if value is None else value * cpi[2024] / cpi[year],
            }
    return records

def window(years, state, event):
    keep = {t: v["y"] for t, v in years.items()
            if v["state"] == state and v["y"] is not None and (t < event or event < t <= event + POST_YEARS)}
    pre, post = [t for t in keep if t < event], [t for t in keep if t > event]
    return keep if pre and post else None

def build_stack(treated, records, excluded):
    """treated: list of dicts with npi, state, event, cluster."""
    cells = defaultdict(list)
    for person in treated:
        cells[(person["state"], person["event"])].append(person)
    rows, units_t, units_c = [], {}, {}
    for (state, event), people in sorted(cells.items()):
        cell = f"{state}:{event}"
        for person in people:
            obs = window(records.get(person["npi"], {}), state, event)
            assert obs is not None, f"treated surgeon lacks pre/post data: {person['npi']}"
            units_t[(cell, person["npi"])] = len(obs)
            for t, y in obs.items():
                rows.append((cell, person["npi"], t, y, 1, event, person["cluster"]))
        for npi, years in records.items():
            if npi in excluded:
                continue
            obs = window(years, state, event)
            if obs is None:
                continue
            units_c[(cell, npi)] = len(obs)
            for t, y in obs.items():
                rows.append((cell, npi, t, y, 0, event, "npi:" + npi))
    nt, nc = len(units_t), len(units_c)
    nt_cell, nc_cell = defaultdict(int), defaultdict(int)
    for cell, _ in units_t:
        nt_cell[cell] += 1
    for cell, _ in units_c:
        nc_cell[cell] += 1
    stack = {k: [] for k in ("cell", "npi", "year", "y", "treated", "event_time", "cluster", "weight")}
    for cell, npi, t, y, d, event, cluster in rows:
        assert nc_cell[cell] > 0, f"cell without controls: {cell}"
        weight = 1.0 if d else (nt_cell[cell] / nt) / (nc_cell[cell] / nc)
        for key, value in zip(stack, (cell, npi, t, y, d, t - event, cluster, weight)):
            stack[key].append(value)
```

```
    stack = {k: np.array(v) for k, v in stack.items()}
    stack["counts"] = {"treated_surgeons": nt, "control_cell_units": nc,
                       "distinct_controls": len({n for _, n in units_c}), "cells": len(cells),
                       "observations": int(len(stack["y"]))}
    return stack

def codes(values):
    _, inverse = np.unique(values, return_inverse=True)
    return inverse

def absorb(matrix, groups, weight, tol=1e-11, max_iter=10000):
    """Weighted residuals of each column on additive fixed effects (alternating projections)."""
    out = np.array(matrix, dtype=float, copy=True)
    if out.ndim == 1:
        out = out[:, None]
    sums = [np.bincount(g, weights=weight) for g in groups]
    for _ in range(max_iter):
        delta = 0.0
        for g, s in zip(groups, sums):
            for j in range(out.shape[1]):
                means = np.bincount(g, weights=weight * out[:, j]) / s
                out[:, j] -= means[g]
                delta = max(delta, float(np.max(np.abs(means))))
        if delta < tol * max(1.0, float(np.max(np.abs(out)))):
            return out
    raise RuntimeError("fixed-effect absorption did not converge")

def fit(stack, columns, outcome):
    groups = [codes(np.char.add(np.char.add(stack["cell"].astype(str), "|"), stack["npi"].astype(str))),
              codes(np.char.add(np.char.add(stack["cell"].astype(str), "|"), stack["year"].astype(str)))]
    w = stack["weight"]
    resid = absorb(np.column_stack([outcome] + columns), groups, w)
    y, x = resid[:, 0], resid[:, 1:]
    xtwx = x.T @ (w[:, None] * x)
    beta = np.linalg.solve(xtwx, x.T @ (w * y))
    e = y - x @ beta
    cluster = codes(stack["cluster"])
    scores = np.zeros((cluster.max() + 1, x.shape[1]))
    for j in range(x.shape[1]):
        scores[:, j] = np.bincount(cluster, weights=w * x[:, j] * e, minlength=cluster.max() + 1)
    g = scores.shape[0]
    bread = np.linalg.inv(xtwx)
    vcov = (g / (g - 1)) * bread @ (scores.T @ scores) @ bread
    treated_clusters = len(set(stack["cluster"][stack["treated"] == 1]))
    return {"beta": beta, "se": np.sqrt(np.diag(vcov)), "df": treated_clusters - 1,
            "treated_clusters": treated_clusters, "clusters": g,
            "_y": y, "_x": x[:, 0], "_cluster": cluster, "_w": w, "_c": g / (g - 1)}

def summarize(model, index=0):
    beta, se, df = float(model["beta"][index]), float(model["se"][index]), model["df"]
    crit = stats.t.ppf(0.975, df)
    return {"estimate": beta, "se": se, "df": df, "ci": [beta - crit * se, beta + crit * se],
            "p": float(2 * stats.t.sf(abs(beta / se), df)),
            "treated_clusters": model["treated_clusters"], "clusters": model["clusters"]}

def group_means(stack):
    """Physician-level pre and post mean payment, controls carried at their stack weights."""
    out = {}
    for label, d in (("treated", 1), ("control", 0)):
        units = defaultdict(lambda: {"pre": [], "post": [], "w": 0.0})
        sel = stack["treated"] == d
        for cell, npi, et, y, w in zip(stack["cell"][sel], stack["npi"][sel], stack["event_time"][sel],
                                        stack["y"][sel], stack["weight"][sel]):
            unit = units[(cell, npi)]
            unit["pre" if et < 0 else "post"].append(y)
            unit["w"] = w
        weights = np.array([u["w"] for u in units.values()])
        pre = np.array([np.mean(u["pre"]) for u in units.values()])
        post = np.array([np.mean(u["post"]) for u in units.values()])
        out[label] = {"physicians": len(units), "pre_mean": float(np.average(pre, weights=weights)),
                      "post_mean": float(np.average(post, weights=weights)),
                      "change": float(np.average(post - pre, weights=weights))}
    return out
```

#### *code/publication_longitudinal.py*

```
"""Public algorithm extract. See README and CODE_PROVENANCE.json."""
import time
import numpy as np
from scipy import sparse
import longitudinal_did as did

def fixed_effect_groups(stack):
    return [did.codes(np.char.add(np.char.add(stack["cell"].astype(str), "|"), stack[k].astype(str)))
```

```python
            for k in ("npi", "year")]

class FixedEffects:
    """Exact weighted two-FE projection via unit elimination/year Schur complement.

    Recalculates unit and year nuisance coefficients for EVERY outcome column.
    Disconnected-cell year normal matrices are small; a symmetric pseudoinverse
    handles the one redundant year level in each cell without arbitrary dropping.
    """
    def __init__(self, stack):
        self.w = stack["weight"]
        unit, year = fixed_effect_groups(stack)
        n = len(unit)
        self.u = sparse.csr_matrix((np.ones(n), (unit, np.arange(n))))
        self.t = sparse.csr_matrix((np.ones(n), (year, np.arange(n))))
        self.us = np.asarray(self.u @ self.w)
        self.cross = self.u @ self.t.T.multiply(self.w[:, None])
        normal = (self.t @ self.t.T.multiply(self.w[:, None]) -
                  self.cross.T @ sparse.diags(1 / self.us) @ self.cross).toarray()
        self.inverse = sparse.csr_matrix(np.linalg.pinv(normal, rcond=1e-12, hermitian=True))

    def unit_residual(self, z):
        return z - self.u.T @ ((self.u @ (self.w[:, None] * z)) / self.us[:, None])

    def residual(self, z):
        z = np.asarray(z, dtype=float)
        if z.ndim == 1:
            z = z[:, None]
        z0 = self.unit_residual(z)
        year_coef = self.inverse @ (self.t @ (self.w[:, None] * z0))
        return z0 - self.unit_residual(self.t.T @ year_coef)

def bootstrap_moments(stack, draws=9999, seed=did.SEED, batch_size=64):
    """Compute exact refitted score quadratics, independent of candidate null.

    Under H0 beta=b, restricted residual is M_FE(y-b*d). A wild sample
    b*d + fitted_FE(y-b*d) + v*residual is refitted, giving beta*=b+dy-b*dx.
    sy-b*sx are its unrestricted cluster scores. Unlike the old method, scores
    account for reestimated FE, not simply wild-weighted preabsorbed scores.
    """
    projector = FixedEffects(stack)
    d = ((stack["treated"] == 1) & (stack["event_time"] > 0)).astype(float)
    yx = projector.residual(np.column_stack((stack["y"], d)))
    y, x = yx.T
    w = stack["weight"]
    cl = did.codes(stack["cluster"])
    g = int(cl.max() + 1)
    s = float(x @ (w * x))
    beta = float(x @ (w * y) / s)
    score_operator = sparse.csr_matrix((w * x, (cl, np.arange(len(cl)))), shape=(g, len(cl)))
    observed_scores = score_operator @ (y - beta * x)
    correction = g / (g - 1)
    se = float(np.sqrt(correction * (observed_scores @ observed_scores)) / s)
    rng = np.random.default_rng(seed)
    result = {key: np.empty(draws) for key in ("dy", "dx", "a", "b", "c")}
    started = time.monotonic()
    for start in range(0, draws, batch_size):
        end = min(start + batch_size, draws)
        v = did.WEBB[rng.integers(0, 6, size=(end-start, g))].T
        vy = v[cl] * y[:, None]
        vx = v[cl] * x[:, None]
        projected = projector.residual(np.column_stack((vy, vx)))
        py, px = np.split(projected, 2, axis=1)
        dy, dx = (w * x) @ py / s, (w * x) @ px / s
        sy = score_operator @ (py - x[:, None] * dy)
        sx = score_operator @ (px - x[:, None] * dx)
        for key, value in (("dy", dy), ("dx", dx), ("a", np.sum(sy*sy, axis=0)),
                           ("b", np.sum(sy*sx, axis=0)), ("c", np.sum(sx*sx, axis=0))):
            result[key][start:end] = value
        if start % (batch_size * 20) == 0:
            print(f"bootstrap {end}/{draws}: {time.monotonic()-started:.1f}s", flush=True)
    result.update(beta=beta, se=se, correction=correction, s=s, draws=draws, seed=seed)
    return result

def bootstrap_pvalue(m, null):
    delta = m["dy"] - null * m["dx"]
    variance = m["correction"] * (m["a"] - 2*null*m["b"] + null*null*m["c"]) / m["s"]**2
    tstar = np.abs(delta) / np.sqrt(np.maximum(variance, np.finfo(float).tiny))
    observed = abs((m["beta"] - null) / m["se"])
    # Finite Monte Carlo correction: never export a spuriously exact p=0.
    return float((1 + np.count_nonzero(tstar >= observed)) / (m["draws"] + 1))

def invert_bootstrap(m, alpha=.05, tolerance=.01):
    def bound(direction):
```

```
        inside = m["beta"]
        step = m["se"]
        outside = inside + direction * step
        for _ in range(60):
            if bootstrap_pvalue(m, outside) < alpha:
                break
            step *= 1.5
            outside = m["beta"] + direction * step
        else:
            raise RuntimeError("Cannot bracket bootstrap confidence interval")
        while abs(outside-inside) > tolerance:
            mid = (inside+outside)/2
            if bootstrap_pvalue(m, mid) >= alpha:
                inside = mid
            else:
                outside = mid
        return (inside+outside)/2
    ci = [bound(-1), bound(1)]
    # Inversion may theoretically have disconnected acceptance regions; check
    # a wide regular grid and explicitly disclose any encountered nonmonotonicity.
    grid = np.linspace(m["beta"]-10*m["se"], m["beta"]+10*m["se"], 401)
    accepted = np.array([bootstrap_pvalue(m, x) >= alpha for x in grid])
    transitions = int(np.count_nonzero(np.diff(accepted.astype(int))))
    return {"p": bootstrap_pvalue(m, 0), "ci": ci, "draws": m["draws"], "seed": m["seed"],
            "weights": "Webb six-point", "null_imposed": True,
            "fixed_effects_refit_each_draw": ["cell_by_physician", "cell_by_year"],
            "inversion": "two-sided restricted wild cluster bootstrap t; common draws across nulls",
            "monte_carlo_p": "(1+exceedances)/(draws+1)", "endpoint_tolerance_dollars": tolerance,
            "acceptance_grid_transitions": transitions,
            "acceptance_grid_disconnected": transitions > 2}

def treated_rows(rows):
    return [{"npi": r["npi"], "state": r["state"], "event": int(r["investment_start"][:4]),
             "cluster": "org:" + r["operator_parent"]} for r in rows]

def preinvestment_means(rows, records):
    """Selection statistic uses index-state pre-event outcomes and nothing later."""
    return {r["npi"]: float(np.mean([v["y"] for year, v in records[r["npi"]].items()
                                    if year < int(r["investment_start"][:4]) and v["state"] == r["state"]
                                    and v["y"] is not None])) for r in rows}

def model_pair(stack):
    d = ((stack["treated"] == 1) & (stack["event_time"] > 0)).astype(float)
    result = {"counts": stack["counts"], "levels": did.summarize(did.fit(stack, [d], stack["y"])),
              "log": did.summarize(did.fit(stack, [d], np.log(stack["y"]))) }
    result["log"]["percent"] = float(100*np.expm1(result["log"]["estimate"]))
    result["log"]["percent_ci"] = [float(100*np.expm1(x)) for x in result["log"]["ci"]]
    result["parents"] = sorted(set(stack["cluster"][stack["treated"] == 1]))
    return result

def event_export(surviving):
    labels = {"le_minus4": "≤−4", "minus3": "−3", "minus2": "−2", "plus1": "+1", "plus2": "+2"}
    times = {"le_minus4": -4, "minus3": -3, "minus2": -2, "plus1": 1, "plus2": 2}
    return {arm: {"coefficients": [{"bin": k, "label": labels[k], "event_time": times[k], **v}
                                   for k, v in surviving[arm]["event_study"].items() if k != "reference"],
                  "reference_event_time": -1, "excluded_event_time": 0,
                  "reference_estimated": False, "units": "2024 USD", "log_event_study": None,
                  "log_event_study_status": "not available in surviving JSON; not fabricated",
                  "counts": surviving[arm]["counts"]}
            for arm in ("practice_mso", "operator_owned")}
```

##### *code/smoke_test.py*

```
"""Synthetic algorithm checks; NOT a replication of the clinical study."""
import numpy as np
from scipy import stats
import alternative_tests as tests
import publication_balance as pb
import longitudinal_did as did
import publication_longitudinal as pl


def main():
    a, b = np.array([10., 20., 30., 40.]), np.array([5., 12., 16., 20., 25.])
    assert np.isclose(tests.welch(a, b)["p"], stats.ttest_ind(a, b, equal_var=False).pvalue)
    assert np.isclose(tests.log_ratio(a, b)["ratio"], np.exp(np.log(a).mean() - np.log(b).mean()))
    rows = [{"npi": "synthetic-A", "year": "2020", "group": "connected", "payment_real": "10"},
            {"npi": "synthetic-B", "year": "2020", "group": "comparison", "payment_real": "20"},
            {"npi": "synthetic-B", "year": "2021", "group": "comparison", "payment_real": "30"}]
    summaries = tests.physician_means(rows)
    assert next(r for r in summaries if r["group"] == "comparison")["mean_annual_payment_2024_usd"] == 25
    z = np.array([[0., 0.], [0., 1.], [1., 0.], [1., 1.]])
    weights, _, diag = pb.entropy_weights(z, np.zeros(4, dtype=int), np.array([1.]), np.array([.5, .5]))
    np.testing.assert_allclose(weights, .25)
```

```
    assert diag["max_standardized_balance_error"] < 1e-8
    data = {"ty": np.array([20., 40.]), "cy": np.array([10., 20., 30., 40.]),
            "tz": np.array([[.5, .5], [.5, .5]]), "cz": z,
            "ts": np.zeros(2, dtype=int), "cs": np.zeros(4, dtype=int),
            "person": np.arange(4), "ncontrol": 4, "strata": ["synthetic"]}
    point, _, _, _ = pb.estimate(data)
    assert np.isfinite(point).all()
    rng = np.random.default_rng(7)
    records, treated = {}, []
    for i in range(16):
        key = f"synthetic-{i}"
        d = i < 8
        records[key] = {year: {"state": "XX", "y": 100 + i + 2*(year-2015)
                              + (12 if d and year > 2017 else 0) + rng.normal(0, .1)}
                        for year in range(2015, 2020)}
        if d:
            treated.append({"npi": key, "state": "XX", "event": 2017, "cluster": f"synthetic-parent-{i}"})
    stack = did.build_stack(treated, records, {r["npi"] for r in treated})
    model = pl.model_pair(stack)
    assert abs(model["levels"]["estimate"] - 12) < .2
    projector = pl.FixedEffects(stack)
    resid = projector.residual(np.column_stack([stack["y"]]))
    np.testing.assert_allclose(projector.residual(resid), resid, atol=1e-8)
    moments = pl.bootstrap_moments(stack, draws=19, seed=17, batch_size=8)
    assert 0 < pl.bootstrap_pvalue(moments, 0) <= 1
    print("PASS: Welch, log ratios, physician aggregation, entropy weights, stacked FE, and wild-bootstrap routines.")


if __name__ == "__main__":
    main()
```

#### Appendix A. MANIFEST Checksums

SHA-256 checksums for the analytical files listed in the source package MANIFEST.json. Documentation-file entries and internal packaging identifiers are omitted.


```
{
  "created_utc": "2026-11-09T22:27:41.697432+00:00",
  "content_policy": "Aggregate outputs, organization-level bibliography, analytical algorithms and documentation only.",
  "files": [
    {
      "file": "VALIDATION.json",
      "sha256": "74e1be6c2fe466fa4c7c6f125c36a1b0ada62de56b47a2882fd1f2dbf133cdc6"
    },
    {
      "file": "aggregate_results/TABLE_CONTEXT.txt",
      "sha256": "2f00042c05f136c2859faa4de9d681d31c1a414ed63147019a18014b94598719"
    },
    {
      "file": "aggregate_results/Table_1.csv",
      "sha256": "43ec7c06c5eb89328da286a44568cc3a627e9485209147e793c882e951d22a42"
    },
    {
      "file": "aggregate_results/Table_2.csv",
      "sha256": "c060d24e905ca8c915c0a5d3b28f480be3721e28e28bab558fbdc896c0f3f5c1"
    },
    {
      "file": "aggregate_results/Table_3.csv",
      "sha256": "7674efd1836d4467351c980696677fb0171412b714432358190a8410e8c9b668"
    },
    {
      "file": "aggregate_results/Table_S1.csv",
      "sha256": "5c0fe6db56706207d99cfff5d7a4c3647ce7d8493d81a97e0bd5c99a8f72b5cd"
    },
    {
      "file": "aggregate_results/Table_S2.csv",
      "sha256": "e3b2ee881376622b834df402baa60fdce43b341ed12ee448b3b8e809890a9cea"
    },
    {
      "file": "aggregate_results/Table_S3.csv",
      "sha256": "34987ef03d6c00adfbe37e17f8261b09a8f49a97f633bf077395241ef3353670"
    },
    {
      "file": "aggregate_results/Table_S4.csv",
      "sha256": "a49e95c79538ed2b5e679c5928a4db237f8993c107dd8273c34d493558be587c"
    },
    {
      "file": "aggregate_results/Table_S5.csv",
      "sha256": "9fb1dab2d2605a27cd22147f982994cd3cbcd6e214ed1e6920265bec87bbb653"
    },
    {
      "file": "aggregate_results/Table_S6.csv",
      "sha256": "e35784d51f8a3a6b86c2e3ed9078d03b2d99fdb33e24434a663bf3b5c41e3ffa"
    },
    {
      "file": "aggregate_results/Table_S7.csv",
      "sha256": "b72fa092f82a7fd96e8e37039d69b648a99ebc48fb42b059980786790105665c"
    },
    {
      "file": "aggregate_results/Table_S8.csv",
      "sha256": "d13f1ced293254c6dab7ebf7faa62768a01011db3700c6e5dd4fe39a2463c96c"
    },
    {
      "file": "aggregate_results/Table_S9.csv",
      "sha256": "9867692fbb26e8f3950633f43e53a5d149e26bc9a052edcb962528d27ebe8714"
    },
    {
      "file": "aggregate_results/longitudinal_analysis.json",
      "sha256": "913f1022f4f45de7b6aea13d5b20a508ce7309fa30bce067594df69c433e378c"
    },
    {
      "file": "aggregate_results/weighted_analysis.json",
      "sha256": "df4bb27780f18ff067bfeee363c143fb245d96fd68465b6852a822d3b6bd18bc"
    },
    {
      "file": "code/CODE_PROVENANCE.json",
      "sha256": "87030db82f119edc1399d002bf53a23edc5f9a4721ba49dc55a7db674d3d5765"
    },
    {
      "file": "code/INPUT_SCHEMA.txt",
      "sha256": "c0a006aef58c9423b6c7e1be875a2c4641b236abea4247a05c92a4803e8b5618"
    },
    {
      "file": "code/alternative_tests.py",
```

```
      "sha256": "ec80642d723ab80c94b5cfe1428733fd320644d7e9228be93ba1351023092602"
    },
    {
      "file": "code/longitudinal_did.py",
      "sha256": "6dea5b4aebdc91160dcd99520fa71560875a2f7a135735879901b7efa865ea8d"
    },
    {
      "file": "code/positive_balance.py",
      "sha256": "9f4119bf4d219b7281423b8dfc6b0b562566a482529237da3064bc2bdc141f68"
    },
    {
      "file": "code/primary_analysis.py",
      "sha256": "bbad9d56d5f04fd6014a6d87dd7c4023ae9df4c927dae02cf42c657b8ac8c3ca"
    },
    {
      "file": "code/publication_balance.py",
      "sha256": "652d0e74ab11ea1fc6e219e8d3219215e5c6dab64cf064f7580818eeb2519c5e"
    },
    {
      "file": "code/publication_longitudinal.py",
      "sha256": "194f18aec3552c5812a762e2580b19ea1380178834fe57cbe57be09d159d947a"
    },
    {
      "file": "code/requirements.txt",
      "sha256": "247cac82bee90f66c969c1966d2ed2e953e746037593bba521fe878b0c84aa40"
    },
    {
      "file": "code/run_analysis.py",
      "sha256": "def89055c238a1c5ff848739a6adbed7cd978efff22620452051d1b3fd46cfb5"
    },
    {
      "file": "code/smoke_test.py",
      "sha256": "7f12f170ec71dd6a0a57963e2fee99bc5ca992fae385079c00ecd8f6d4266b25"
    },
    {
      "file": "investment_bibliography/README.txt",
      "sha256": "28632a14bdcb4c9067b5993570e9f92f706d5abcd803e5cf646be0d1e9aff3f9"
    },
    {
      "file": "investment_bibliography/organization_windows.csv",
      "sha256": "5c0fe6db56706207d99cfff5d7a4c3647ce7d8493d81a97e0bd5c99a8f72b5cd"
    },
    {
      "file": "investment_bibliography/sources.csv",
      "sha256": "053bfa7c6b75a23b9c641f6b416c6c17b98545e45d433f6e6ddc77224abe0d3a"
    },
    {
      "file": "timing_audit/README.txt",
      "sha256": "6cef102504e5be48a4af8f92fb9b63ae9aeb089c53f82d1742b5aefc5641861a"
    },
    {
      "file": "timing_audit/aggregate_timing_audit.json",
      "sha256": "47006b19d8c7e68acad3468fd892dc7b7d49f3ae59078a3c5425f65d50dbe87f"
    }
  ]
}
```
